# Data sharing policies, requirements, and support from public and private clinical trial sponsors: a survey on top sponsors of clinical trials in Europe

**DOI:** 10.64898/2026.03.31.26349853

**Authors:** Ka Hin Tai, Giulia Varvarà, Emma Escoffier, Ulrich Mansman, Nicholas J. DeVito, Anna Catharina V. Armond, Florian Naudet

## Abstract

**Objective:** To map the presence, public availability, and content of clinical trial data sharing policies (DSP), data management and sharing plans (DMSP), and data use agreements (DUA) among the most prolific public and private clinical trial sponsors operating in the European Union, and to identify key areas of convergence, divergence, and constraint in the context of General Data Protection Regulation (GDPR).

**Eligibility criteria:** We included organisation-level documents describing approaches to clinical trial data sharing or data management from the top 20 public and top 20 private sponsors ranked by the number of trials registered in the EU Clinical Trials Information System (CTIS). Eligible materials comprised publicly available or sponsor-shared policies, guidelines, statements, templates, and agreements relevant to clinical trial data sharing or management.

**Sources of evidence:** Evidence was identified through systematic searches of sponsors’ public websites, structured Google searches, and major data management plan platforms (DMPTool, DMPonline, DMP Assistant), complemented by direct contact with sponsors to verify findings and request missing documentation. All sources were archived and catalogued.

**Charting methods:** Two reviewers independently extracted data using a structured form, capturing the existence, accessibility, and content of data sharing policies, data management and sharing plans, and data use agreements. Quantitative data were summarised descriptively, and a non-interpretive descriptive content analysis was conducted to characterise recurring policy elements and areas of heterogeneity.

**Results:** Among 40 sponsors, private sponsors were substantially more likely than public sponsors to make trial-specific data sharing policies and data use agreements publicly accessible, often via established data sharing platforms. Public sponsors more frequently referenced data management and sharing plans, but these were heterogeneous in scope and often embedded within broader institutional governance documents rather than tailored to clinical trials. Across sectors, GDPR compliance, data protection, and legal safeguards were emphasised, while operational aspects such as dataset readiness, review criteria, and downstream responsibilities varied widely. Overall response rate to sponsor verification was 37.5%.

**Conclusion:** Clinical trial data sharing governance in the EU shows a marked sectoral imbalance among the top sponsors. Private sponsors tend to provide more detailed and operationally explicit documentation, whereas public sponsors often articulate high-level commitments without trial-specific guidance. Greater clarity and standardisation, particularly among public sponsors, could improve transparency and facilitate responsible data reuse, while remaining compatible with GDPR requirements.

## Introduction

The sharing of clinical trial data has gained increasing recognition as a pillar for transparent, reproducible and ethical research, as it can reduce research waste and fulfill the ethical obligation toward the patients^1^. Many steps have recently been taken by journals, funders, and regulators to improve data sharing^1–4^. This includes their efforts to encourage, and in some cases require, research sponsors to set in place robust data sharing and data management policies, especially those supporting clinical trials. Sponsor policies often ensure compliance with these newer data sharing requirements, as well as define the research environment within their institution.

Sharing clinical trial data tends to encounter more difficulties than other research data, as it contains personal data from the patients^5^. In the European Union (EU), the sharing of personal data is regulated by the General Data Protection Regulation (GDPR)^6^, which contains both opportunities and constraints for responsible data sharing. All health data is defined as sensitive under the GDPR; therefore, it can be processed only after having obtained formal consent from the subjects, and its processing needs to be rigorously controlled and tracked. Data controllers, a defined role under GDPR, now have to clearly describe which data is being processed for which purpose and by whom. They also have to track all transfers or processing of said data and assess all the possible associated risks^7^.

All these measures allow reduction in the risk of data misuse and give the subjects more control over their privacy, but they also hinder clinical trials’ data sharing, as compliance with the GDPR requires stricter data protection measures. These measures increase research and data management costs, requiring additional time and infrastructure from data providers. As the GDPR applies to every subject in the EU, private and public sponsors of trials operating in Europe must therefore adapt to this regulatory context, which presents specific challenges. Despite the increased focus on transparent clinical trial data, it is unclear to what extent the sponsors in the European Union have developed and made publicly available, well-defined, and transparent data sharing policies.

This study intends to complement a recent scoping review on data sharing and data management plans. Here, we applied a narrower focus, concentrating only on the sponsors of clinical trials acting in Europe and choosing the top 20, based on clinical trial activity, from both the private and the public sector. We aim to use this sample to understand the presence and public availability of data sharing policies and agreements among leading sponsors and their common elements. This will identify the most relevant areas of concern, most common restrictions, and any requirements to obtain data access. Similarly, we also collected information on data management requirements and the presence of data management plans from each of the sponsors, to obtain an overview of the characteristics of data management and its level of standardization across sponsors.

## Methods

### Protocol and Registration

This survey was conducted in accordance with the CARDA guidance for document analyses (Checklist for Assessment and Reporting of Document Analysis)^8^. The study protocol was registered on the Open Science Framework prior to data collection (https://osf.io/93cbv).

### Identification and Selection of Sponsors

Following the protocol, we identified the top 20 public and top 20 private sponsors by the number of registered clinical trials in the EU Clinical Trials Information System (CTIS). Public sponsors were defined as non-commercial or public entities, where private sponsors were defined as private, for-profit entities, most commonly pharmaceutical companies. Sponsor names were extracted and normalised through fuzzy-matching (Python 3.12.2, fuzzywuzzy package), with iterative manual review of potential duplicates or variant spellings. Separate ranked lists for academic and commercial institutions were generated, and the top 20 sponsors in each category were selected for assessment.

### Information Sources and Search Strategy

We examined publicly available organisational documentation relevant to clinical trial data sharing and data management. Eligible materials included policies, guidelines, statements, templates, data use agreements, and any other documents describing institutional approaches to clinical trial data sharing or data management. Documents were included regardless of format (e.g., webpages, PDFs, policy statements, legally-oriented documents) so long as they covered organisational-level procedures. For each included organisation, we systematically searched publicly available sources using (1) on-site navigation and native search tools, and (2) a structured set of general search terms applied to both the organisation’s website and to the first three pages of Google search result. Search terms included: *“data sharing”*, *“clinical trials”*, *“data management plan”*, *“data sharing plan”*, and *“data governance”, along with the sponsor name*. Searchers were permitted to adapt or expand terms as needed, and additional terms identified inductively during the review were applied retrospectively across all organisations.

For non-English materials, we used a combination of native speakers and machine translation tools (Google Translate, DeepL). We also screened major Data Management Plan platforms (DMPTool, DMPonline, DMP Assistant) to identify any publicly posted DMPs related to the included organisations.

All webpages and documents were downloaded, archived, and catalogued.

### Data Collection and Extraction

Each organisation was independently assessed by two reviewers (KH, GV, EE), who extracted data using a structured Google Form designed specifically for the study. Extracted elements included: Existence and type of data sharing policy; verbatim text of relevant policies; conditions or exceptions for sharing; documentation related to data management and sharing plans; existence and content of structured templates; presence and contents of data use agreements; participation in DORA and CoARA (academic sponsors only); mentioning of the PhRMA-EFPIA Principles for Responsible Clinical Trial Data Sharing (private sponsors only); and existence of data champion programs.

Extraction was performed independently. Disagreements were resolved through discussion between the two reviewers or by consulting a third senior team member (FN) when necessary. For each funder, information was extracted from all eligible documents and aggregated inclusively, whereby any item reported in at least one document was retained. When discrepancies arose across documents, the funder was contacted for clarification. All documents were stored in structured folders on Google Drive.

In accordance with the protocol, we identified at least one public contact per organisation (e.g., Data Protection Officer, research administration, data governance representative, public relationship or media). Organisations were contacted to verify extracted information and invited to provide missing documents, clarifications, or corrections. All correspondence was logged. Institutions that did not respond were followed up at 1-week and 3-week intervals and were given up to 4 weeks to reply. Any additional evidence submitted, whether publicly available or shared privately, was assessed and incorporated, with confidentiality of individuals preserved.

### Quantitative Analysis

Quantitative data were summarised descriptively using counts and percentages. Analyses and visualisations were performed in R Version 4.4.1 (RRID:SCR_001905), following transparent and reproducible coding practices.

### Qualitative Analysis

A descriptive content analysis was conducted to characterise the components of data sharing policies, data management and sharing plans, and data use agreements. Consistent with the protocol, this analysis adopted a non-interpretive, manifest document-analysis approach^9^ and did not impose theoretical frameworks.

Two researchers independently reviewed an initial sample of documents and jointly developed draft codebooks for each document type. After iterative piloting and refinement, the coding frameworks were applied across all included materials. Coding disagreements were resolved through discussion. Extracted qualitative data were organised into structured lists reflecting recurring policy elements and areas of divergence. Coding and data extraction were conducted manually using shared documents and spreadsheets (Google Docs and Google Sheets) to record coded elements and supporting excerpts.

### Open Science and Data Availability

All extracted data, archived documentation, correspondence records (with identifying details removed), and analytic scripts were deposited on the Open Science Framework (https://osf.io/3kc2r/) in accordance with the open science commitments of this study.

### Deviation from original protocol

During post hoc verification of sponsor identification and ranking using CTIS data, we found that some public sponsors were listed under multiple name variants and were not fully merged during the initial matching process. As a result, one public sponsor that met the initial inclusion threshold was not included, and another sponsor near the cutoff was included instead. This deviation is described further in the Limitations.

## Results

### Identification of Sponsors and Data Collection

The full EU Clinical Trials Information System (CTIS) dataset from inception to July 2025, which contained 9,481 registered trials, was downloaded on 3 July 2025 using the platform’s built-in “Download results” function. Supplementary figure 1 displays the sponsor selection and document assessment process. Across this dataset, 1,996 unique sponsors were identified, including 705 public and 1,291 private sponsors, accounting for 4,286 and 5,195 registered trials, respectively. We identified the top 20 public and top 20 private sponsors by the number of registered clinical trials in the CTIS. Supplementary table 1 shows the full list of the included sponsors. As Celgene (ranked 14 on the list) was acquired by BMS in 2019 and data sharing policy or any other online material was no longer available, we replaced it by the next in rank, Sun Pharma. Supplementary figure 2 shows the distribution of registered clinical trials by sponsor rank for public and private sponsors, with summary counts and the top-20 inclusion threshold indicated. Beyond the top-20 threshold, trial activity among public sponsors was highly dispersed. The 20 included public sponsors accounted for 1,244 of 4,286 publicly sponsored trials (29.0%), with the remaining 3,054 trials (71.0%) distributed across 685 additional public sponsors. In comparison, the 20 included private sponsors accounted for 2,106 of 5,195 privately sponsored trials (40.5%), with the remaining 3,181 trials (59.5%) distributed across 1,271 additional private sponsors. This indicates that while both sectors exhibit dispersion beyond the highest-volume sponsors, trial activity in the public sector appears more structurally fragmented.

The included industry sponsors in total have more registered trials than the public sponsors (n = 2106 trials compared to n = 1244). The distribution among private sponsors was more right-skewed (mean 105.3 vs median 92.5), whereas public sponsors showed a more symmetric distribution (mean 62.2, median 62.2). The interquartile range was substantially larger among private sponsors (85.5 vs 26.0), suggesting greater concentration among a subset of large contributors. All included industry sponsors are pharmaceutical companies, whereas the public sponsors are a mix of organisations (1 university, 3 public hospitals, 11 university hospitals, 3 research organisations, 2 municipal authorities). Geographically, six public sponsors came from the Netherlands, five from France, three from Denmark, two from Sweden, and one each from Austria, Norway, Belgium, and Italy. Three public sponsors or their directly affiliated institutes signed DORA and six signed CoARA, while Novartis was the only private sponsor who signed on to DORA. Bristol-Myers Squibb and three public sponsors offered data champion programs, which trained researchers to better manage research data. Table 1 shows the characteristics of the included sponsors.

**Table 1.**
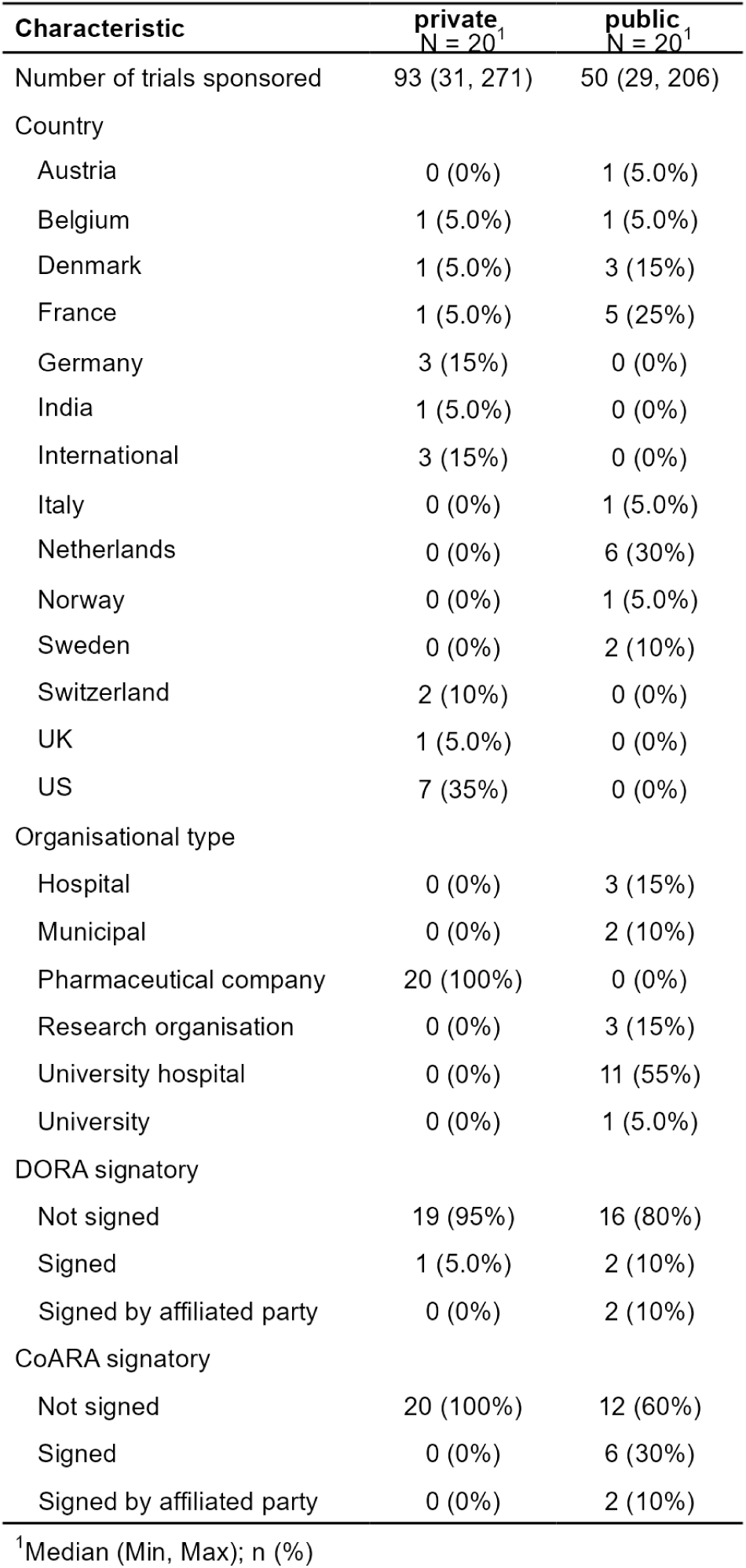
Characteristics of the 40 included sponsors.

#### Box 1 Documents examined in this study

Data Sharing Policy (DSP) - A policy defining whether, when, and under what conditions clinical trial data may be shared, typically set at the level of the data holder or responsible organisation.

Data Management and Sharing Plan (DMSP) - A trial-specific document describing how clinical trial data will be generated, curated, protected, and prepared for sharing, including timelines, access routes, and safeguards for participant confidentiality.

Data Use Agreement (DUA) - A legally binding contract governing secondary use of shared clinical trial data, specifying permitted analyses, data-security obligations, publication rights, and restrictions on re-identification.

We collected data in August and September 2025 on sponsors’ data sharing policies (DSPs), data management and sharing plans (DMSPs), and data use agreements (DUAs) from their public-facing websites. Many of the private sponsors have their data sharing details described on trial data sharing platforms (17 on Vivli, 2 on CSDR, 1 on YODA). Three major data management plan platforms (DMPtool, DMPonline, and DMPassistant) were queried but no additional generic information about the sponsors was found (yet we identified some individual projects from the public sponsors posting on those platforms). Supplementary table 2 lists all the websites and online material of sponsors we abstracted data from. Many documents from public sponsors were written in their local languages. Translation tools like DeepL were used to support data extraction.

The extracted findings were sent to the sponsors’ representatives for confirmation. Nine private sponsors and six public sponsors responded, who either verified or amended our findings, and some provided additional documents for our reference. Total response rate was 37.5% and median response time was 16 days. At the initiative of the Open Science officers at Ospedale San Raffaele, a virtual call was held to further support verification of the extracted data.

Figure 1 shows the number of sponsors with their DSP, DMSP, or DUA identified and assessed in this study. Many private sponsors posted their DSPs on data sharing platforms, and adapted the DUA templates from the platforms. Numerous public sponsors openly mentioned that they actively engaged in sharing research data, adapted the FAIR principles, and supported open science. However, they did not always detail their policy on data sharing, and thus we counted them as sponsors with identified but not assessed material. The existence of DMSP was seldom confirmed by private sponsors, whereas the DMSP could usually be found as a part of research integrity guidelines among public sponsors.

**Figure 1.**
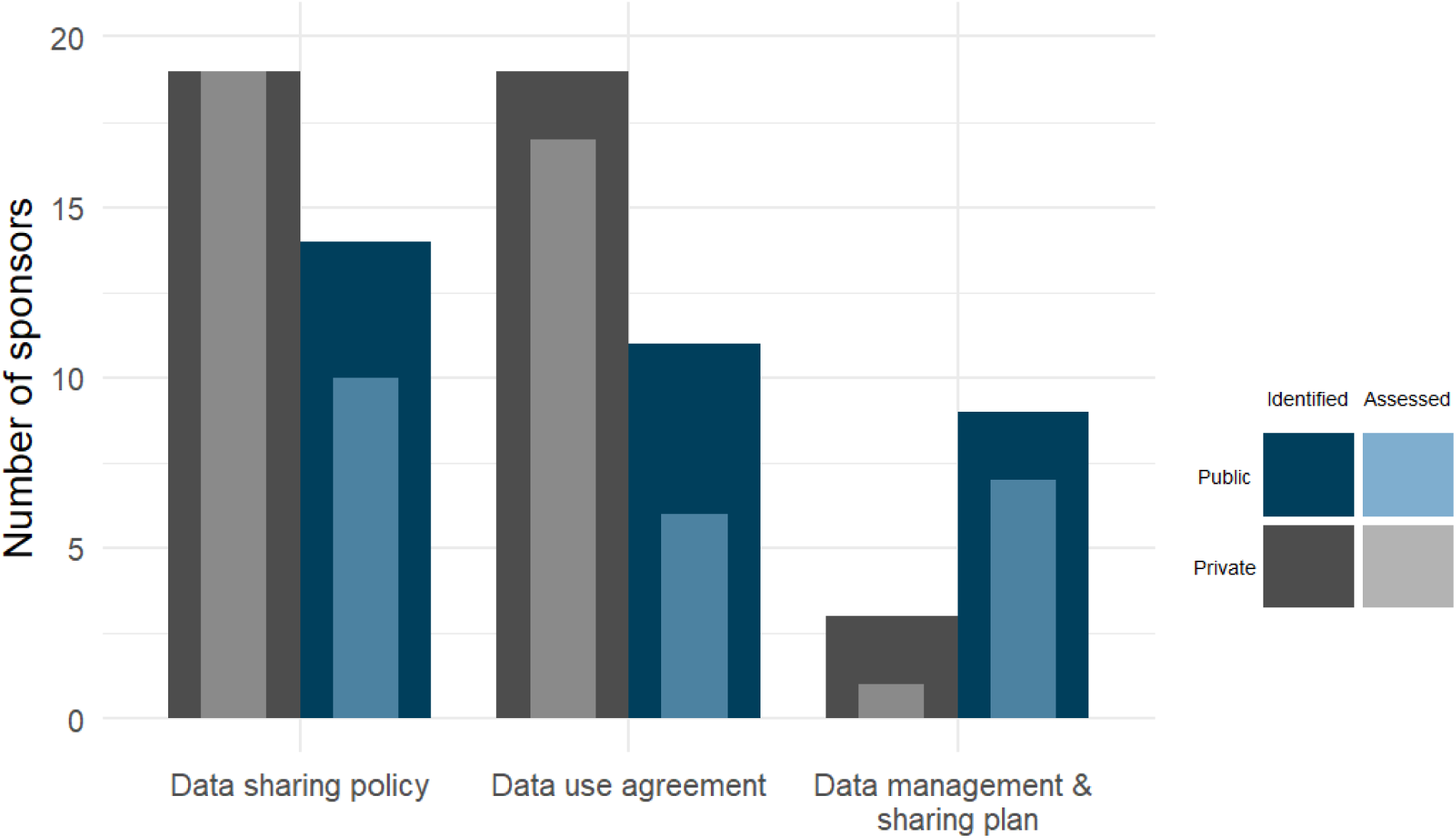
Number of documents identified and assessed for analysis. *The “assessed” documents were the materials that were accessible to the study team and could be directly reviewed, either because they were publicly available or otherwise shared with us by the sponsors. The “identified-only” documents were the materials whose existence could be confirmed but that were not accessible for review in this study. These documents were either mentioned or referenced in other sources, or admitted by sponsors in our correspondence as restricted, internal documents*.

### Data Sharing Policy (DSP)

We examined 19 DSPs from private sponsors and 10 from public sponsors. Novo Nordisk upgraded its data sharing framework during this study and we extracted the updated information. The majority of private sponsors (n = 17) used a data sharing platform to facilitate their data sharing activities, and two sponsors accepted data sharing requests via email. Merck Sharp Dohme developed its own data sharing request portal and shared their data for approved projects via Vivli. Unlike private sponsors, which commonly relied on established data-sharing platforms, public sponsors more often developed or adopted institution-specific or regional repositories. For example, Radboud UMC hosted data via the Radboud Data Repository, while the Netherlands Cancer Institute used the Health Data Space Amsterdam. UniCancer, together with Gustave Roussy, developed the WeShare infrastructure. Odense University Hospital relied on the OPEN Network, whereas Utrecht UMC, Amsterdam UMC, and Erasmus MC shared data via the DataverseNL platform. Finally, Ospedale San Raffaele established its own Open Research Data Repository for data storage and sharing.

Differences were observed between private and public sponsors in the availability and clarity of their data sharing policies. The private sponsors (pharmaceutical companies) were in general trial-oriented, of which 13 sponsors were signatories to the PhRMA-EFPIA Principles for Responsible Clinical Trial Data Sharing. They openly documented their data sharing policies with detailed information, to enhance clinical trial transparency and data sharing. For the identified public sponsors, on the other hand, despite their substantial contribution to clinical research and their large share of CTIS-registered trials, they were institutions with a broad research mandate, not primarily dedicated to conducting clinical trials. Thus, less trial-relevant information about data sharing could be extracted from the public sponsors. In addition, the orientation of the documents differed markedly between sectors. Policies from private sponsors were largely outward-facing, with a primary focus on guiding external researchers through the process of requesting access to trial data. In contrast, documents from public sponsors were more inward-facing, often framed as guidance for investigators and staff on how to manage, prepare, and share data, rather than as instructions for prospective data requestors.

Figure 2 displays the frequency of various items about clinical trial data sharing found in the DSPs among private sponsors. The items were categorised into five domains: namely general data sharing policy, types of data shared, data sharing restrictions, data anonymisation, and data request and review process. The emphasis placed on specific data sharing elements varied across the five domains. While private sponsors showed broadly similar patterns in the types of data shared and data request review processes, their approaches to anonymisation and the restrictions applied were much heterogeneous. For example, elements such as the research objective and hypothesis, the analysis plan and publication plan, and relevant legal or contractual considerations were commonly deemed necessary when evaluating data access requests. In contrast, expectations regarding the research funding of the request teams, dataset readiness and quality, and the practicality of the proposed project varied more widely.

**Figure 2.**
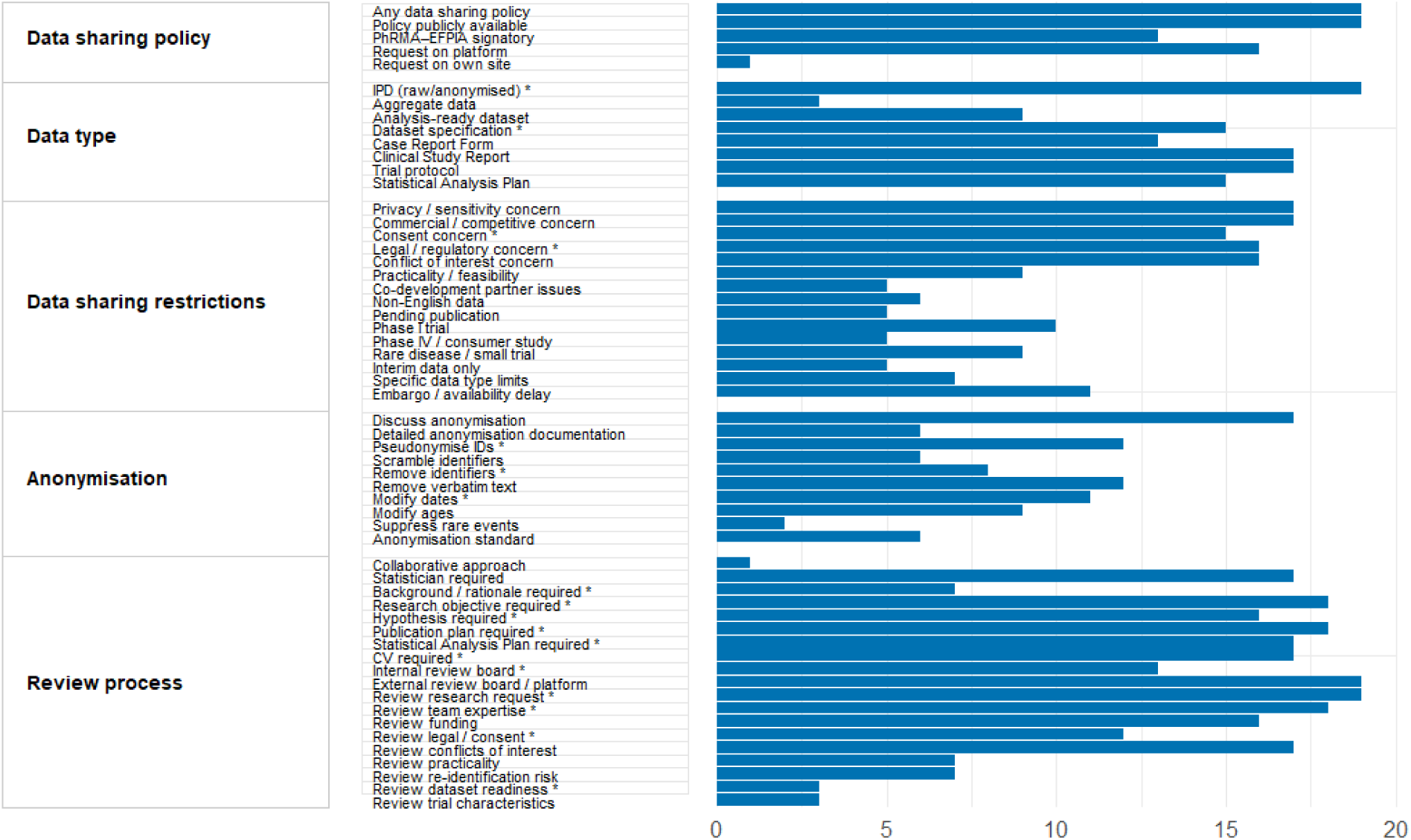
Content of clinical trial data sharing policy among private sponsors. *\* Items marked with an asterisk were also identified among sponsors, but only infrequently.*

Note that 17 of the private sponsors posted structured data sharing details on Vivli. Vivli, and other similar platforms, imposed a common set of requirements and standardised data request pipeline, thus the findings among those sponsors were more homogeneous compared to the disparate platforms used by others. Yet, the sponsors could still choose to emphasise certain elements from a list of defaults, or provide additional information as needed. Our analysis captured both sponsor-specific details and data sharing platform-specific norms.

In contrast to private sponsors, publicly available information on clinical trial data sharing among public sponsors was sparse and heterogeneous. Only ten public sponsors articulated any form of data sharing policy, and these policies were typically framed at the level of general research data rather than being tailored specifically to clinical trial data. With respect to the types of data shared, four public sponsors explicitly mentioned the possibility of sharing patient-level data. However, references to supplementary trial materials, such as trial protocols, statistical analysis plans, or case report forms, were rare or absent. Information on data sharing restrictions was even more limited. Only two public sponsors discussed legal considerations or conflicts of interest as part of their data sharing conditions, and other common restrictions observed among private sponsors were largely undocumented in the public sector. Finally, data request and review processes were seldom described in detail. Among the public sponsors examined, only Rigshospitalet and UMC Utrecht provided substantive descriptions of their review procedures, outlining criteria for assessing whether a data request would be considered qualified. Overall, the limited scope and trial-specificity of publicly available policies precluded a systematic graphical comparison with private sponsors.

### Data Management and Sharing Plan (DMSP) or Template

Data management and sharing plans were scarce among private sponsors. Boehringer Ingelheim was the only private sponsor with a publicly-available document that resembled a DMSP, while two other private sponsors reported the existence of internal DMSP documents that were not accessible to us. In distinction to private sponsors, DMSPs were more frequently identified among public sponsors (n=9), where they were occasionally embedded within broader institutional research integrity or governance guidelines. Several university hospitals actively used the DMPonline platform, registering project-specific DMSPs in the repository. Although these project-specific DMSPs reflected research practice, they were excluded from our analysis as they were not generic clinical trial data sharing templates. Individual study teams were responsible for the development of their own DMSPs and the resultant documents usually ended up different from each other. However, on the research support page of UMC Utrecht, we found that the sponsor had DMP templates registered on the DMPonline platform (not publicly available), where researchers at UMC Utrecht could use and adapt the templates for their own projects.

Supplementary table 3 outlines numerous domains and items identified in the DMSPs from the seven public sponsors. While all sponsors addressed core components, such as governance, data protection, storage, documentation, and data sharing, the depth and operational specificity varied substantially. Data protection and legal compliance were described in the greatest detail, with extensive guidance on GDPR obligations, risk assessments, handling of identifiable or sensitive data, and safeguards against data breaches. In contrast, upstream elements such as cost attribution, data quality control, and practical reuse of existing data were less consistently operationalised. Although long-term storage, retention, and repository deposition were commonly mandated, expectations for reproducibility, metadata standardisation, and data availability timelines were articulated with varying clarity.

### Data Use Agreement (DUA)

While 19 privates and 11 public sponsors explicitly mentioned the implementation of data use agreement during their data sharing request process, only 17 and 6 sponsors, respectively, made their DUA templates publicly available. Among the private sponsors, 16 adopted the DUA template from Vivli, one from CSDR, one from YODA, and two drafted their own DUAs. GlaxoSmithKline had its data hosted on both Vivli and CSDR, but since 2023 all data requests were directed to the Vivli platform, presumably only the Vivli DUA was then applicable. Novartis joined Vivli in December 2025, and currently its data can be requested through either Vivli or CSDR. All the 11 public sponsors either devised their own DUA or adopted regional or national templates.

The DUAs could be arranged in five parts: Scope and general provision, Data use conditions, Publication requirements, Intellectual property terms, and Legal terms and conditions. Since private sponsors utilised data sharing platforms to process data requests, they ought to adopt the DUA templates from the platforms. Sponsors using the same data sharing platform were required to adopt a common DUA template and therefore presented similar elements. Although sponsors may tailor these templates on a case-by-case basis, we did not have access to the final, modified DUAs.

Figure 3 illustrates substantial heterogeneity in coverage and emphasis of data use agreement terms among private and public sponsors. Core agreement items such as data use restrictions, data privacy protections, legal and regulatory compliance, and publication requirements were consistently addressed by nearly all sponsors. In contrast, some items were less uniformly covered, especially among public sponsors. Divergence was most pronounced for provisions related to operational constraints (e.g. restricted parties, notification of covered person status) and dataset readiness (e.g. guarantee of data accuracy). Intellectual property and publication-related clauses were widely included across both sectors, whereas discussion related to downstream legal terms showed great variability.

**Figure 3.**
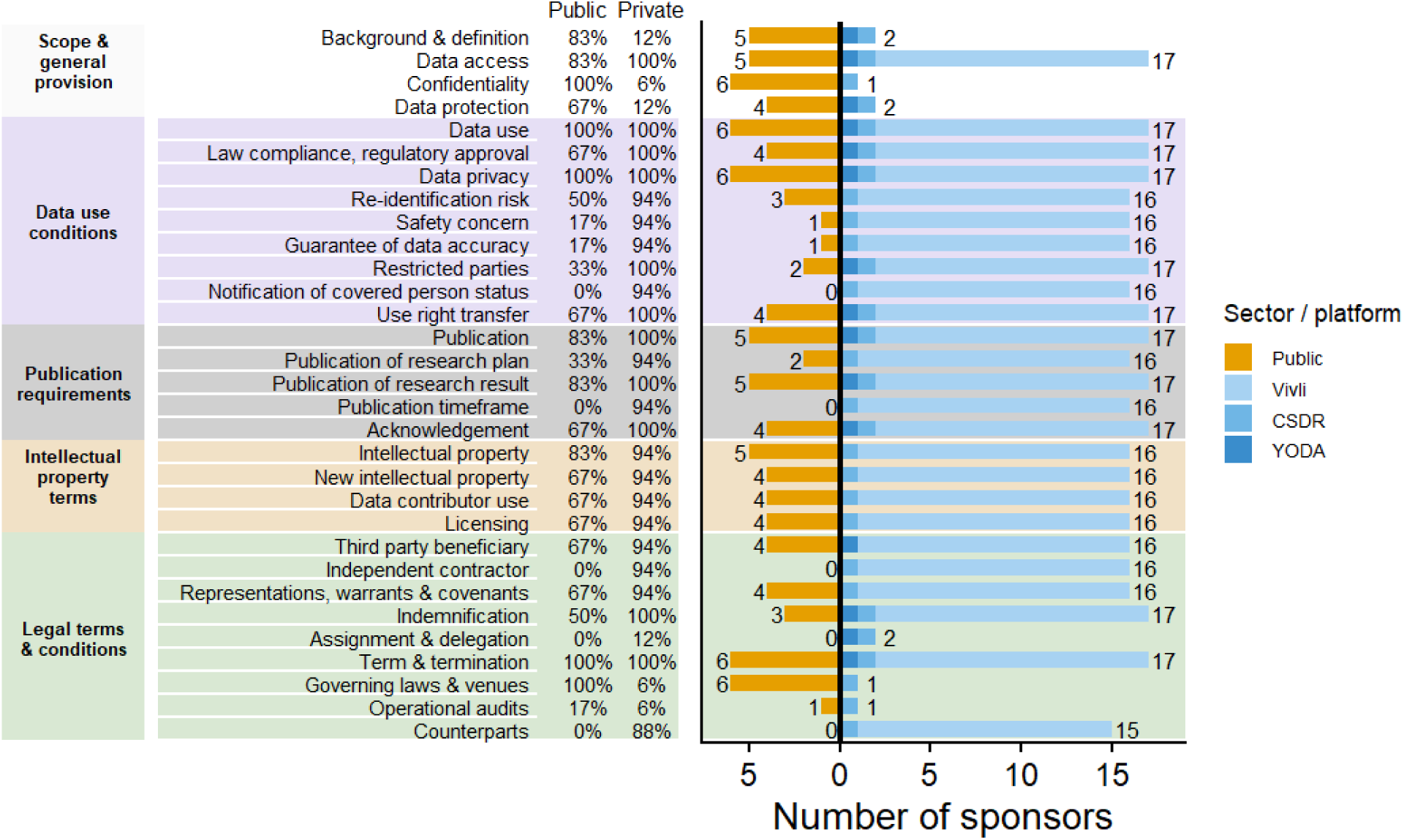
Comparison of data use agreement content between private and public sponsors. *Percentages indicate the proportion of sponsors mentioning each item among those who made a DUA publicly available.*

Incyte Corp. and Gilead Sciences Inc. both referenced the use of a DUA in their own data sharing policies. However, one sponsor indicated that the DUA was an internal document and therefore not shared, while the other did not respond to our request and no DUA could be identified online. Consequently, neither was summarised in Figure 3. Available information from their public websites, nonetheless, suggested that data use agreements considered data privacy, risk of patient re-identification, and legal compliance, alongside with publication conditions and intellectual property arrangements. UniCancer was developing WeShare, a national digital research infrastructure, with scientific coordination by Gustave Roussy. The initiative aimed to establish a health research data repository alongside a suite of digital research tools. According to the consortium’s operating charter, a data use contract governing data access, data use, publication, and data protection was being implemented.

## Discussion

In this survey of 40 of the most prolific clinical trial sponsors registered in the EU Clinical Trials Information System, we observed a high degree of heterogeneity in how clinical trial data sharing is governed, documented, and communicated. Substantial variation was evident in the availability, specificity, and operationalisation of data sharing policies, data management and sharing plans, and data use agreements.

Within this diverse landscape, certain patterns were associated with sponsor type. Private sponsors, which in our sample consisted of pharmaceutical companies, more often made trial-specific data sharing policies and data use agreements publicly accessible, often via established data sharing platforms. These documents tended to be detailed, standardised, and focused on downstream data access governance, including eligibility criteria, review processes, and legal safeguards. In contrast, public sponsors commonly articulated high-level commitments to open science and FAIR principles. They more often relied on broader institutional documentation, with less consistently trial-specific, publicly accessible documentation detailing how clinical trial data could be requested, assessed, and shared in practice. Data management and sharing plans were more commonly identifiable among public sponsors than private sponsors, but these documents varied widely in scope and operational detail and were often embedded within broader institutional research governance frameworks rather than tailored to clinical trials.

### Findings in Context

These findings closely mirror those reported in a 2020 survey of commercial and non-commercial therapeutic research funders, which showed that non-commercial funders primarily emphasised upstream requirements such as data management planning and adherence to FAIR principles, whereas commercial funders focused more explicitly on downstream mechanisms for requesting and accessing individual participant data^10^. Audits of pharmaceutical company transparency policies have similarly found that many large industry sponsors have formal commitments to sharing individual patient data, although the scope and clarity of these commitments vary considerably across companies^11^. Consistent with this broader literature, our findings suggest that differences in emphasis across sponsor types remain evident. Public, non-commercial funders were less likely to rely on established clinical data-sharing platforms (e.g. Vivli, YODA), and more often addressed data sharing through broader institutional or project-level documentation. Explicit references to individual participant data sharing and formalised processes for reviewing data access requests were also less frequently described in publicly available materials.

Normative and theoretical work has long emphasised that effective clinical trial data sharing requires more than aspirational commitments. In a seminal framework, Mello et al. argued that responsible data sharing systems should balance broad access to participant-level data with accountability to both data providers and data requesters, while remaining practical and sustainable to operate^12^. Central to this framework is the need for transparency, clearly defined roles, and explicit governance mechanisms, such as articulated access criteria, review processes, and enforceable data use conditions, to ensure that data sharing is both trustworthy and workable in practice^12^. More recently, Modi et al. translated these principles into concrete recommendations for data sharing policy design, calling for publicly accessible, clearly communicated, and regularly updated policies that specify what data are shared, when they are shared, under what conditions, and through which processes, supported by supplementary documentation such as trial listings, protocols, and data access materials^13^. When assessed against these theoretical and policy-oriented benchmarks, our findings suggest uneven alignment between recommended governance principles and their operationalisation in current sponsor-level documentation. While high-level commitments to transparency and open science were frequently articulated among public sponsors, detailed descriptions of data access pathways, review mechanisms, and data use conditions were often fragmented across documents.

Prior studies suggest that limitations in clinical trial data sharing emerge at multiple points along the data sharing pipeline. At the trial level, non-industry-funded studies are less likely to declare an intention to share data at registration, and stated plans frequently change between registration and publication, highlighting the fragility of upstream signalling alone^14^. At the funder level, non-commercial funders often acknowledge the importance of IPD sharing without mandating it, relying instead on supportive or aspirational policy language^15^. In contrast, commercial funders more frequently articulate formal IPD sharing policies^16^, yet empirical evaluations demonstrate that actual data access remains limited and heterogeneous, with substantial barriers to eligibility, delays in provision, and variable utility of shared IPD packages^17,18^. Our findings occupy the intermediate governance layer of this pipeline, showing that public sponsors differ from private sponsors in both trial-level signalling and policy specificity, and less frequently operationalise data sharing through platforms, review processes, and data use agreements, which are the critical structures for translating intent and policy into meaningful data access.

One interpretation is that private sponsors, despite operating under stronger commercial and legal constraints, have faced sustained external pressure from regulators, journals, funders, and civil society, to formalise and standardise their clinical trial data sharing practices. This pressure may have incentivised investment in dedicated infrastructures and legally robust documentation. Public sponsors, by contrast, often operate within complex institutional environments with broader research mandates, decentralised governance structures, and heterogeneous legal responsibilities. As a result, clinical trial data sharing may be addressed indirectly through general data governance or research integrity policies, rather than through dedicated, trial-oriented frameworks. While such approaches may offer flexibility to individual researchers, they risk creating ambiguity for external researchers seeking access to clinical trial data. Recent reporting guidance, including the updated CONSORT 2025^19^ and SPIRIT 2025^20^ statements, emphasises the need for structured data sharing plans and clear description of access procedures. In practice, however, many trial reports still rely on generic statements such as “data available upon reasonable request”, which provide limited operational guidance on how requests are evaluated or implemented^21^. In decentralised institutional environments, this also creates a risk that data sharing arrangements are not institutionally preserved over time, as they may rely on individual staff members rather than on formalised systems.

The heterogeneity observed in data use agreements further underscores this tension. Although core elements, such as data protection, legal compliance, and publication conditions, were consistently addressed across sectors, divergence emerged for provisions related to operational feasibility, dataset readiness, and downstream responsibilities. These variations may affect the likelihood of data access being granted, as well as the efficiency, reproducibility, and interpretability of secondary research using shared trial data. Together, these findings suggest that greater alignment in data use agreement content could help better accommodate the needs of the research community while preserving necessary legal and ethical safeguards.

Sponsor identification and ranking were based on publicly available CTIS registry data, which required harmonisation of sponsor names that were variably recorded across submissions. Although automated and manual checks were applied, post hoc verification identified that several public sponsors were registered under multiple name variants that were not fully merged during the initial ranking process. This led to the omission of one otherwise eligible public sponsor from the final top-20 selection and the inclusion of another sponsor whose trial count was close to the inclusion threshold. Importantly, this affected only a single sponsor at the margin of the public sponsor ranking and did not alter the overall distribution of documents, the patterns of heterogeneity observed, or the study’s conclusions. The primary impact is a partial loss of qualitative information from the omitted sponsor, rather than a systematic bias in the comparative analysis.

### Strengths and Limitations

A key strength of this study is the systematic and transparent mapping of sponsor-level documentation across both public and private sectors, using a predefined protocol and dual independent extraction. Importantly, we actively contacted sponsors to verify findings and invite corrections or supplementary materials, allowing organisations an opportunity to respond and reducing the risk of misclassification based solely on public-facing information. The inclusion of both quantitative summaries and descriptive content analysis provides a nuanced overview of not only whether documents exist, but how they are structured and what they prioritise.

Several limitations should be acknowledged. First, our focus on the top 40 sponsors by trial volume limits generalisability. As shown in Supplementary Figure 2, the top 20 sponsors in each sector accounted for only a minority of registered trials (28.8% public; 38.8% private), indicating that a substantial proportion of trial activity lies outside the highest-volume institutions. Smaller sponsors or those operating primarily outside the EU regulatory context may exhibit different practices. Second, although CTIS provided a centralised regulatory registry of clinical trials taking place in Europe, it primarily focused on interventional clinical trials with an Investigational Medicinal Product, typically new medications or indications submitted as part of marketing authorisation or label extension. Thus, sponsors, such as University Hospital of Munich or Charité University Hospital Berlin, who predominantly conducted treatment-optimisation trials using already market-approved medicinal products, were not required to register such trials on CTIS and hence are likely under-represented in our study. Third, reliance on publicly accessible documents inevitably introduces availability bias. Internal policies, bespoke agreements, or case-specific modifications, which were particularly common among public sponsors, were often inaccessible and therefore could not be assessed. Fourth, our analysis captures a snapshot in time. Sponsor policies are evolving rapidly, especially in response to regulatory changes, technological developments, and growing expectations around data reuse. In fact, Novo Nordisk and Novartis updated their data sharing policies in December 2025 and we updated our findings accordingly. Finally, we did not evaluate how policies are implemented in practice or how frequently data requests are approved, which limits inference about the real-world impact of documented policies.

Despite these limitations, our findings have several implications. First, they highlight a need for greater clarity and standardisation in how public sponsors articulate clinical trial data sharing expectations. Even relatively lightweight, trial-specific guidance, covering scope, request pathways, review criteria, and responsibilities, could substantially improve transparency without imposing undue administrative burden. Second, this work provides an empirical baseline against which future developments can be assessed. As initiatives promoting FAIRification, data altruism, and secondary use of health data mature, longitudinal analyses will be essential to track whether policy convergence occurs across sectors and whether increased formalisation leads to greater data reuse. Finally, our findings are particularly relevant in the European context. The strong emphasis on GDPR compliance observed across documents reflects legitimate concerns around data protection and re-identification risk. However, variability in how these concerns are operationalised suggests room for clearer, shared interpretations of GDPR-compliant clinical trial data sharing. Harmonised guidance at the European level, bridging regulatory compliance with practical data reuse, could help align expectations across public and private sponsors while safeguarding participant rights.

### Conclusion

In summary, this study reveals a large variety of practices in clinical trial data sharing governance. Addressing the heterogeneity in data sharing policies, data management plans, and data use agreements will require not only normative commitment to openness, but also concrete, transparent, and trial-specific policies that make data sharing feasible in practice within the highly protective regulatory context of the EU General Data Protection Regulation. By mapping current approaches and detailing the various domains identified in each document, this study contributes a foundation for future empirical, normative, and policy-oriented work on improving clinical trial data sharing across sectors.

## Supporting information

Checklist for Assessment and Reporting of Document Analysis (CARDA)

Common items of Data Management and Sharing Plan found from public sponsors

Online material from sponsors assessed for this study

List of included sponsors

## Acknowledgments

This project is a collaboration between the SHARE-CTD doctoral network (funded by the European Union, Marie Curie Grant Agreement Number 101120360) and the EMBRACE project (funded by the French ANR). It also reuses the main structure from an audit of the OSIRIS Consortium (funded by the European Union under the grant agreement No.101094725).

Funded by the European Union. Views and opinions expressed are, however, those of the author(s) only and do not necessarily reflect those of the European Union or the European Commission. Neither the European Union nor the granting authority can be held responsible for them.

## Declaration of interests

KT, GV, UM, and FN received grants provided by SHARE-CTD doctoral network (Horizon-MSCA.2022-DN 101120360). https://share-ctd.eu/ FN also received grants from the French National Research Agency, https://anr.fr/en/, the French Ministry of Health, https://sante.gouv.fr/, the French Ministry of Research, https://www.enseignementsup-recherche.gouv.fr/fr, and the OSIRIS project (Horizon 101094725). https://osiris4r.eu/

## CRediT author contributions

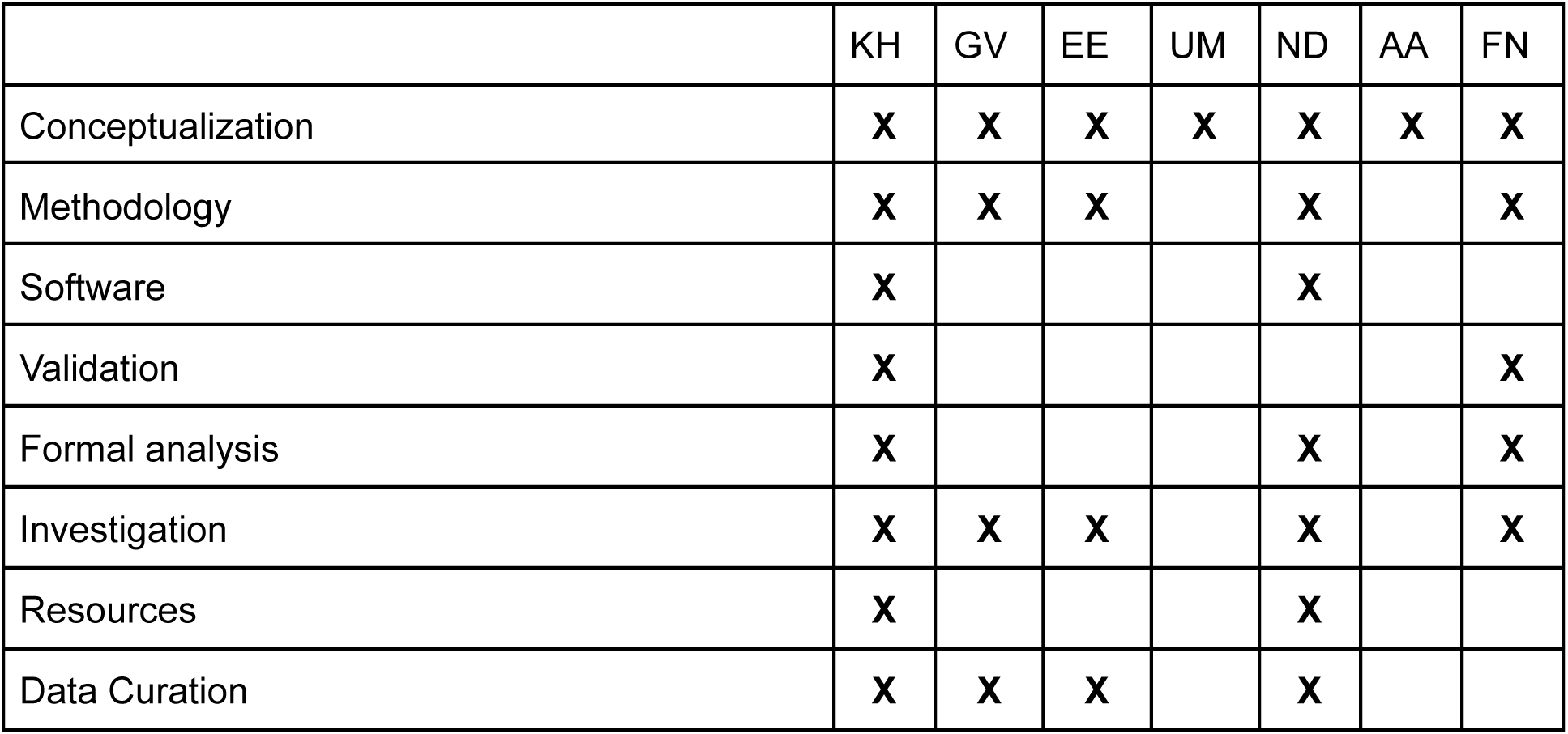
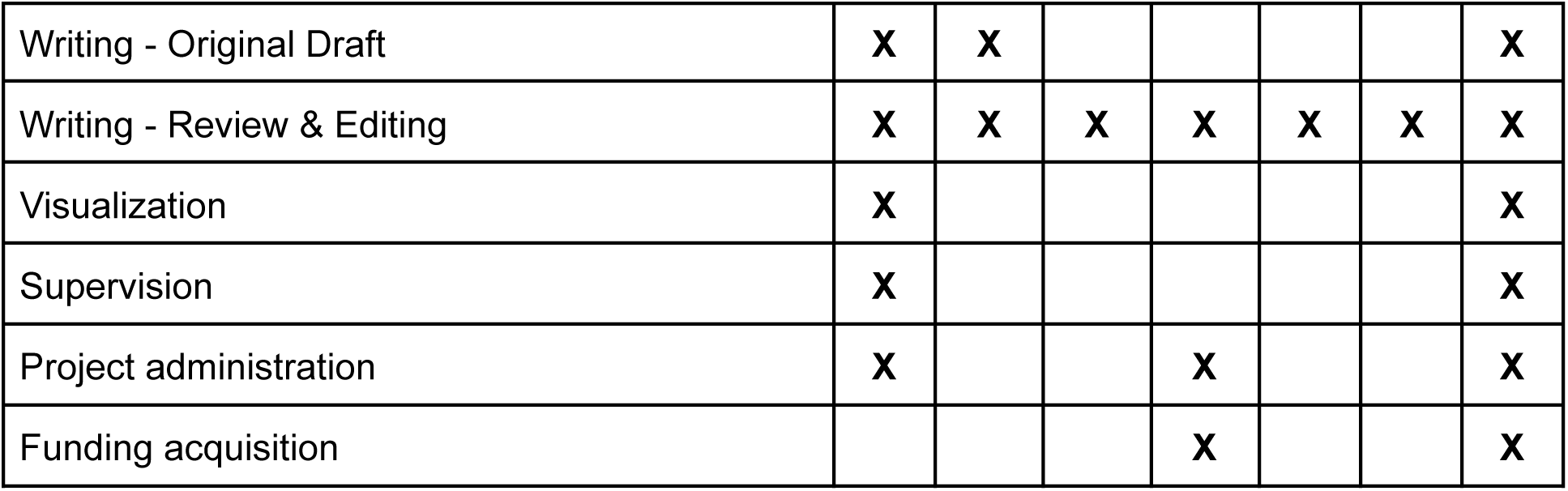

|  | KH | GV | EE | UM | ND | AA | FN |
| --- | --- | --- | --- | --- | --- | --- | --- |
| Conceptualization | X | X | X | X | X | X | X |
| Methodology | X | X | X |  | X |  | X |
| Software | X |  |  |  | X |  |  |
| Validation | X |  |  |  |  |  | X |
| Formal analysis | X |  |  |  | X |  | X |
| Investigation | X | X | X |  | X |  | X |
| Resources | X |  |  |  | X |  |  |
| Data Curation | X | X | X |  | X |  |  |
| Writing - Original Draft | X | X |  |  |  |  | X |
| Writing - Review & Editing | X | X | X | X | X | X | X |
| Visualization | X |  |  |  |  |  | X |
| Supervision | X |  |  |  |  |  | X |
| Project administration | X |  |  | X |  |  | X |
| Funding acquisition |  |  |  | X |  |  | X |

---

**Supplementary figure 1.**
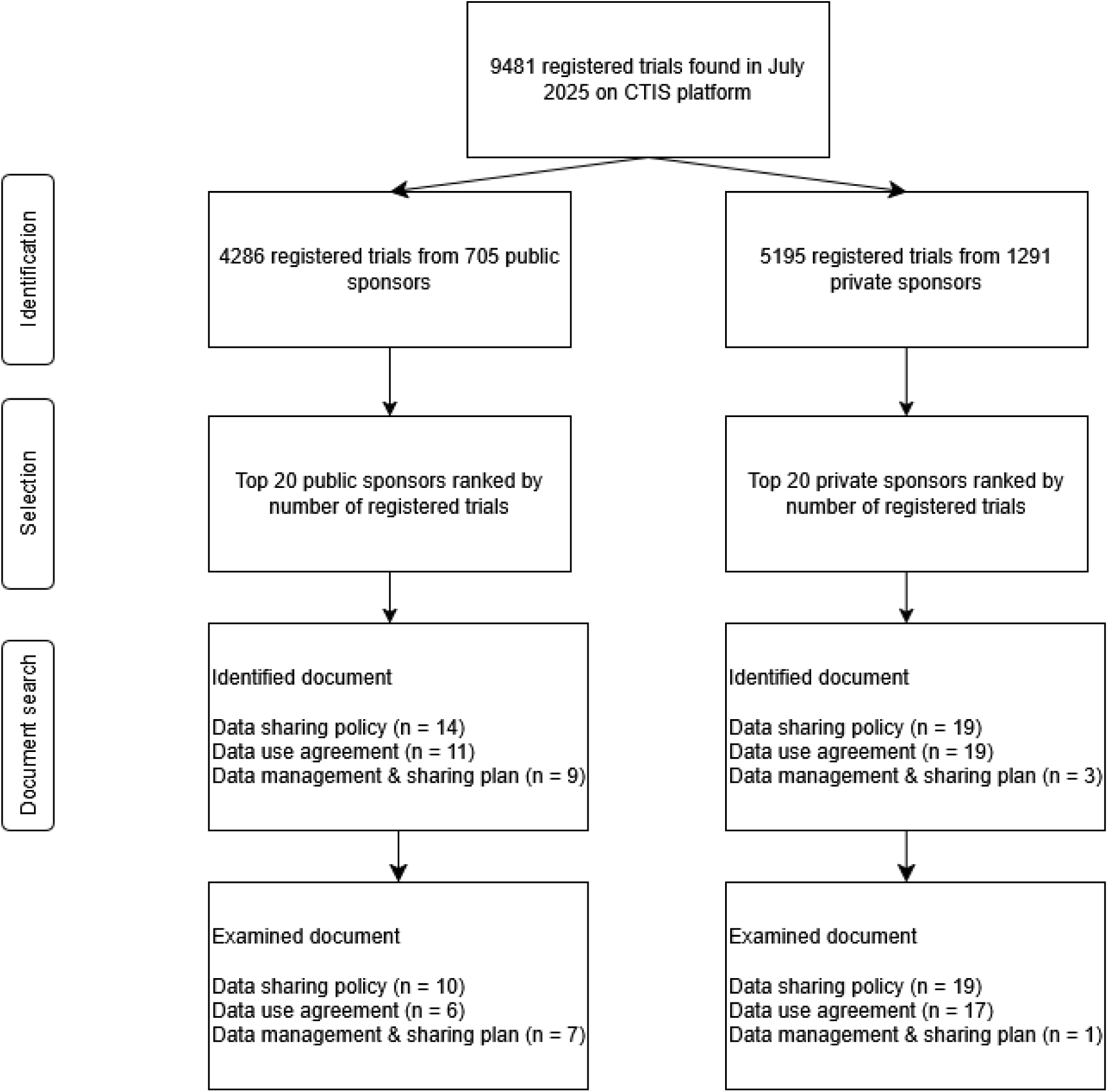
Flow diagram of sponsor identification, selection, and document assessment.

**Supplementary figure 2.**
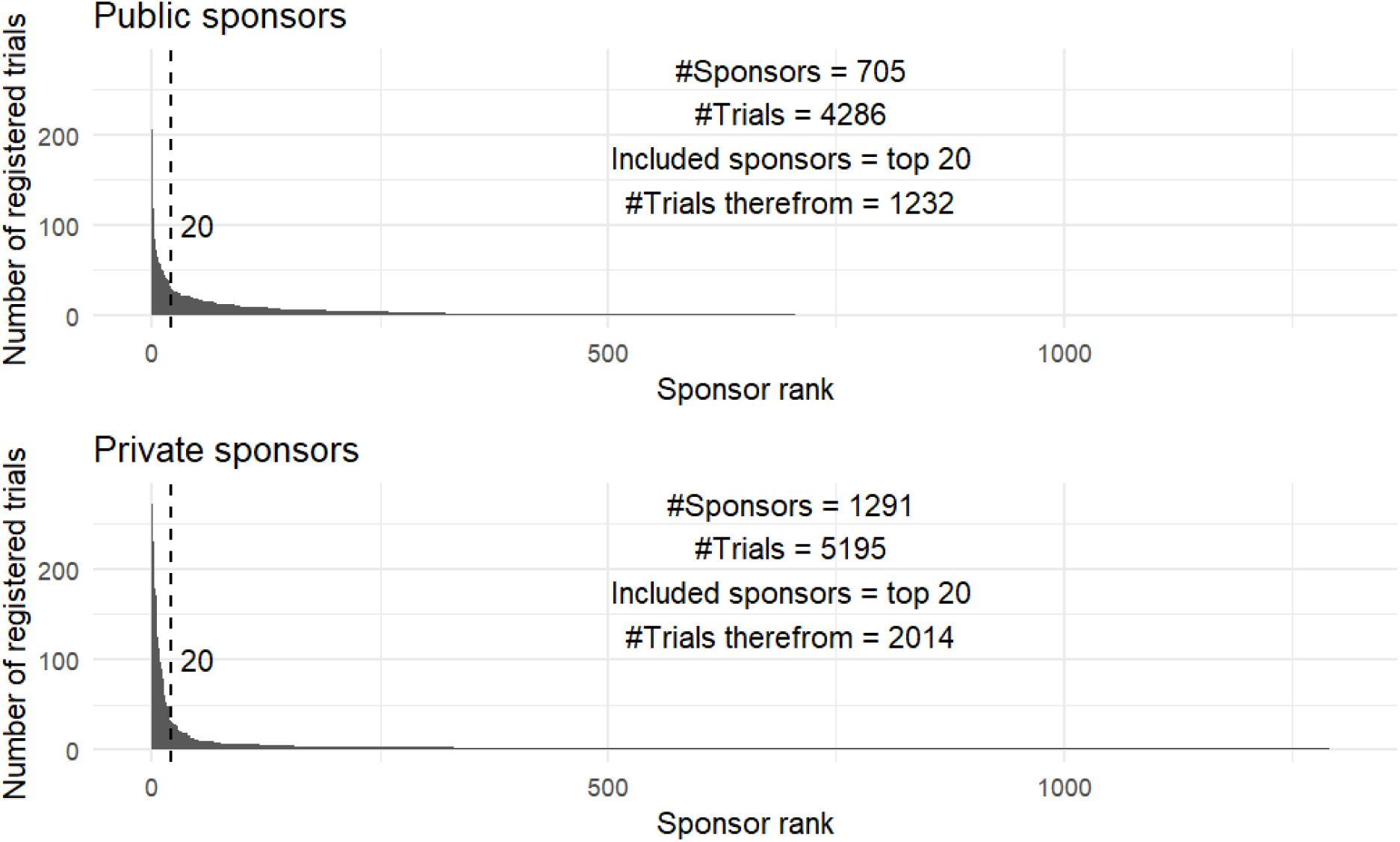
Distribution of number of registered trials by sponsor.

**Supplementary table 1.**
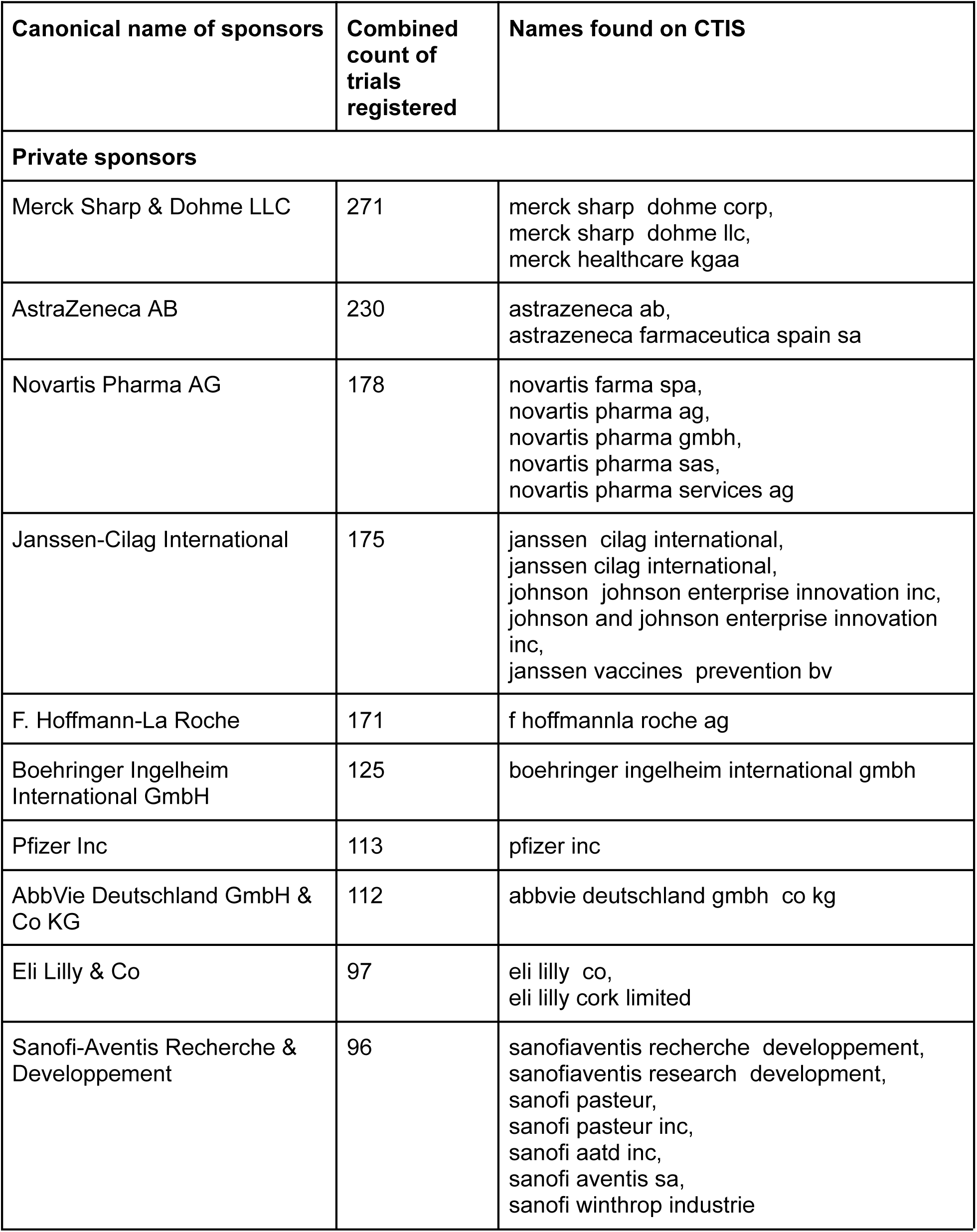
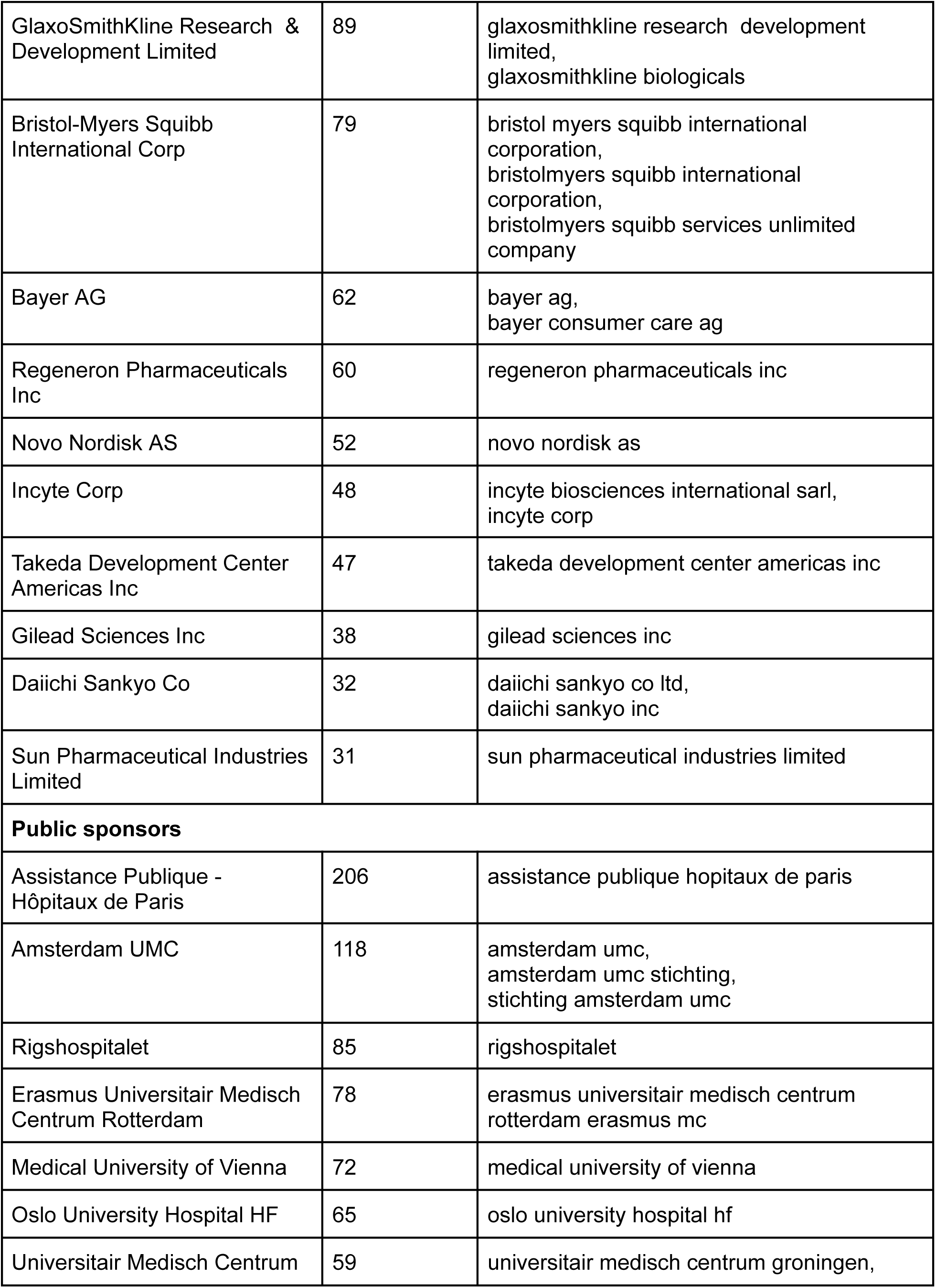
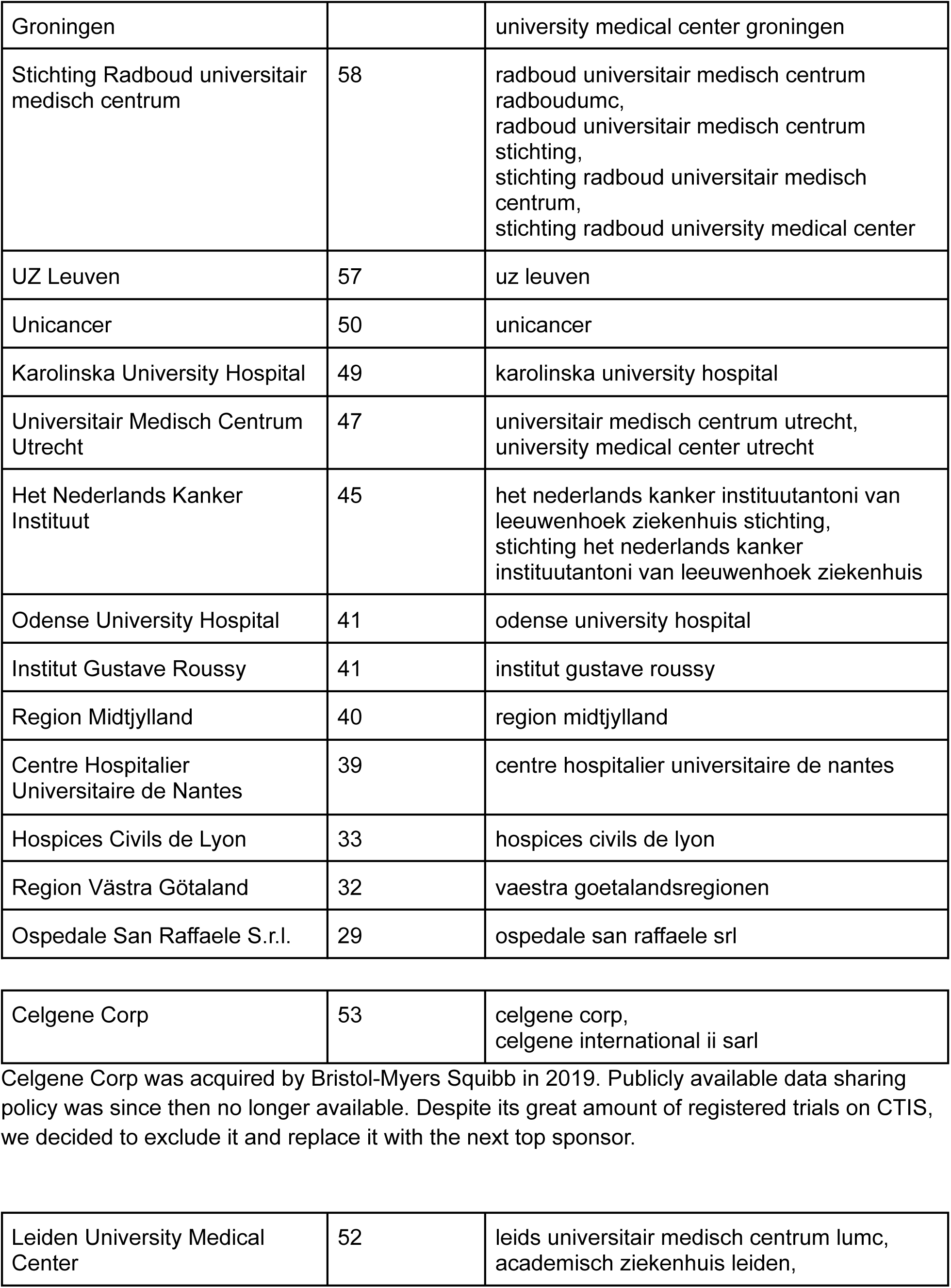
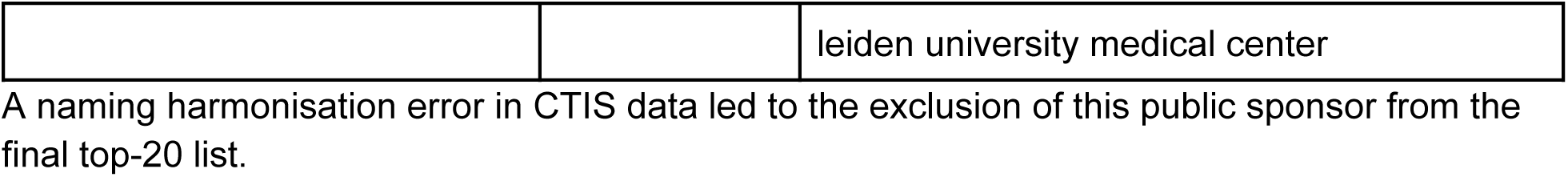
List of included sponsors.

**Supplementary table 2.**
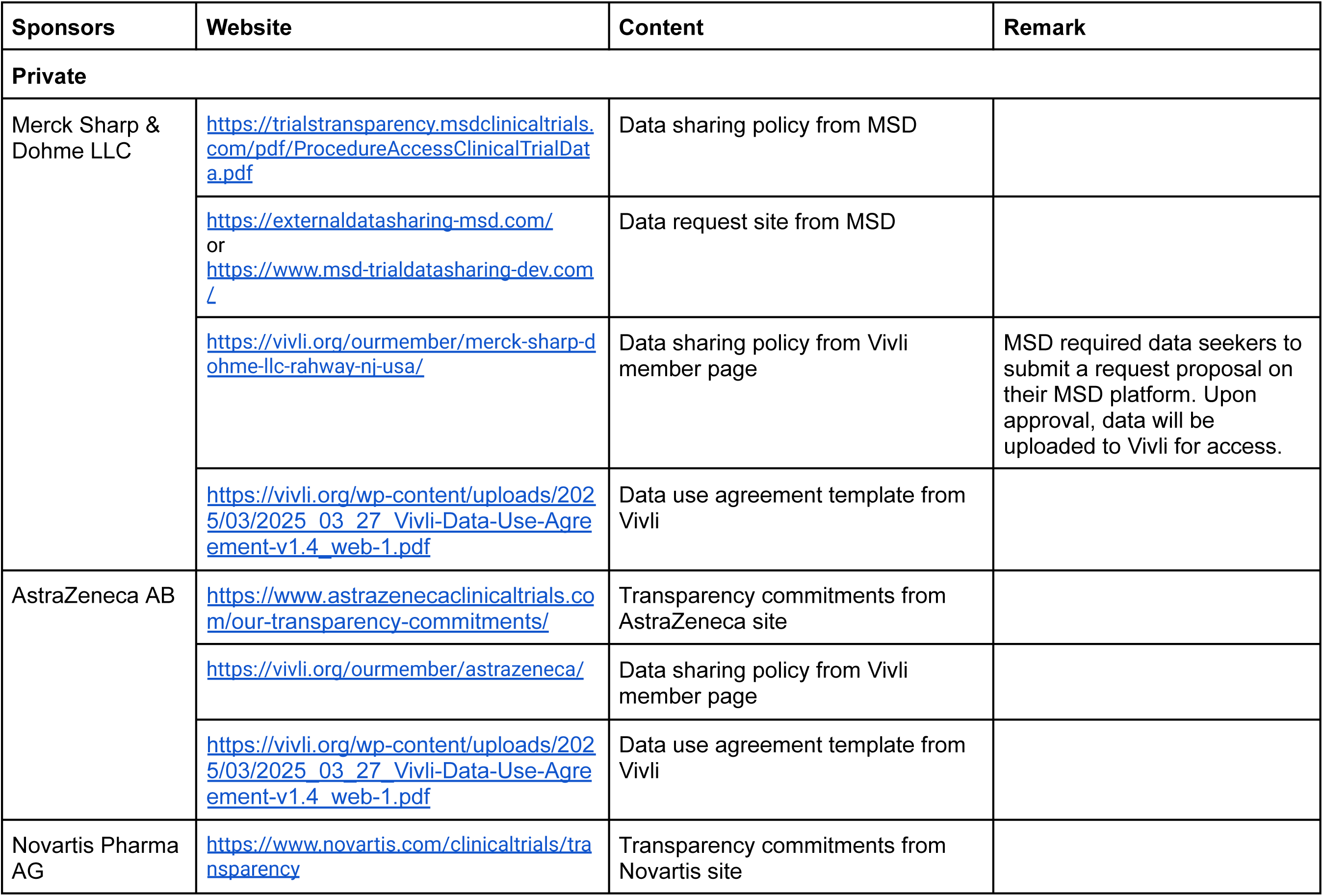
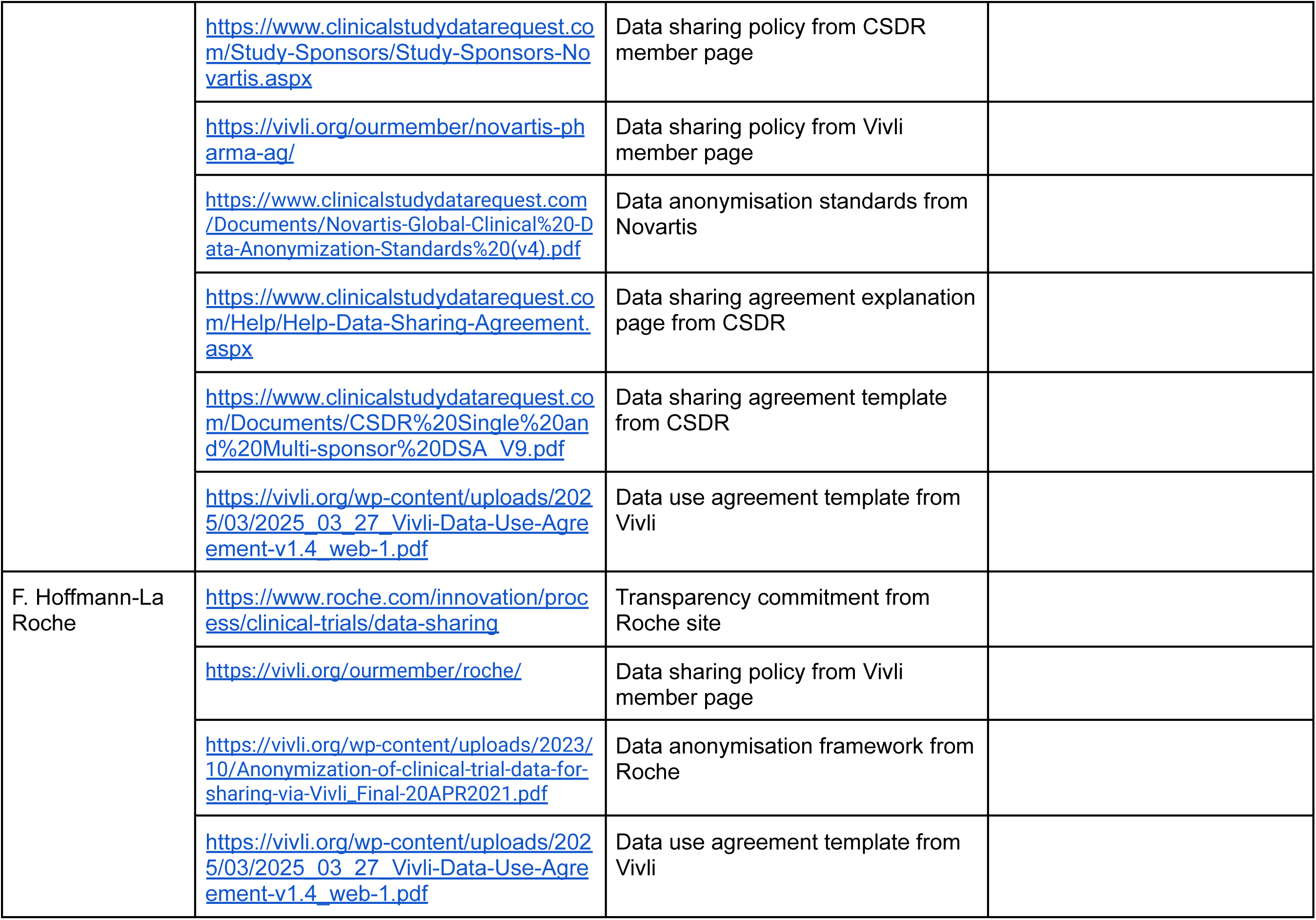
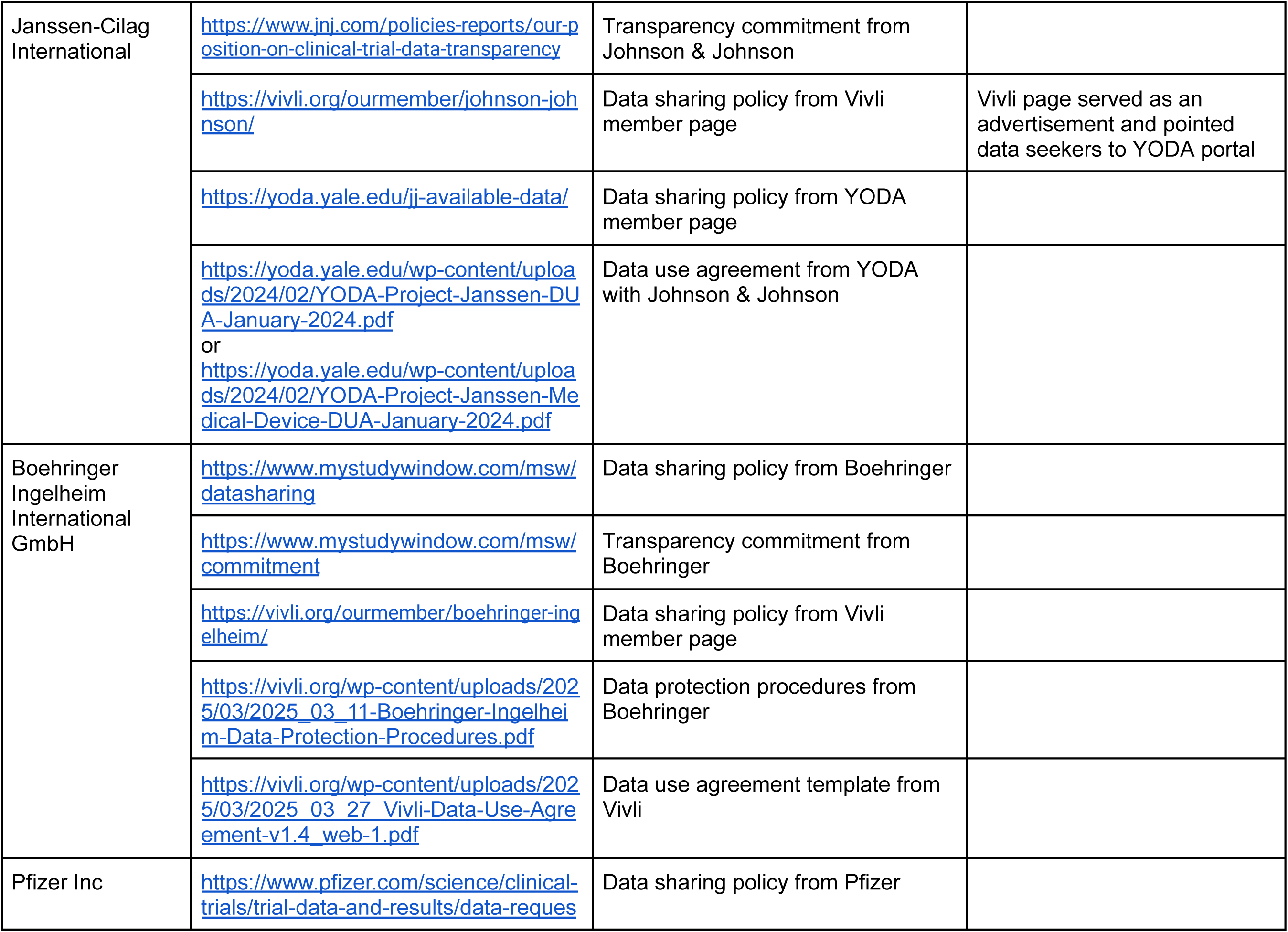
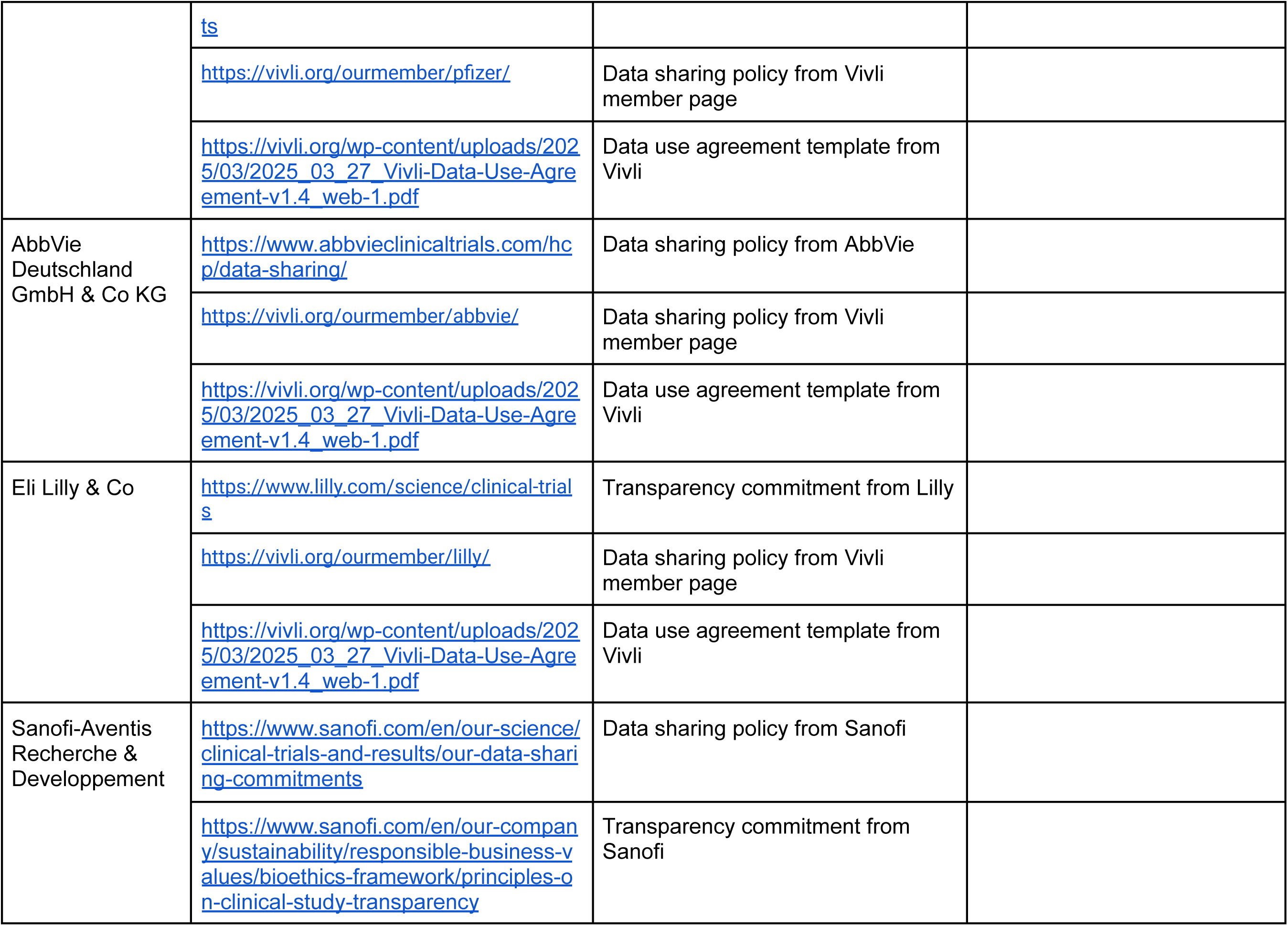
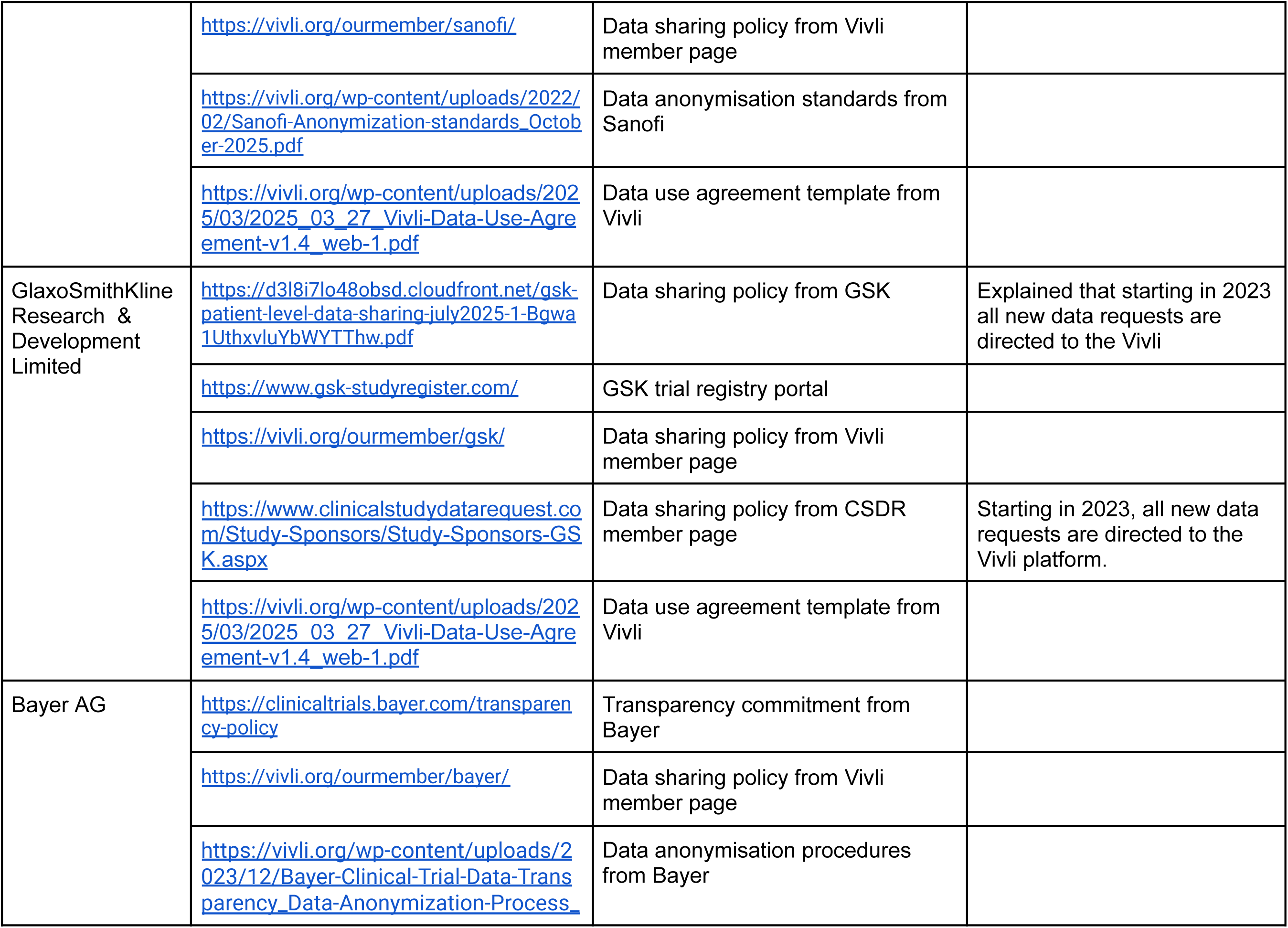
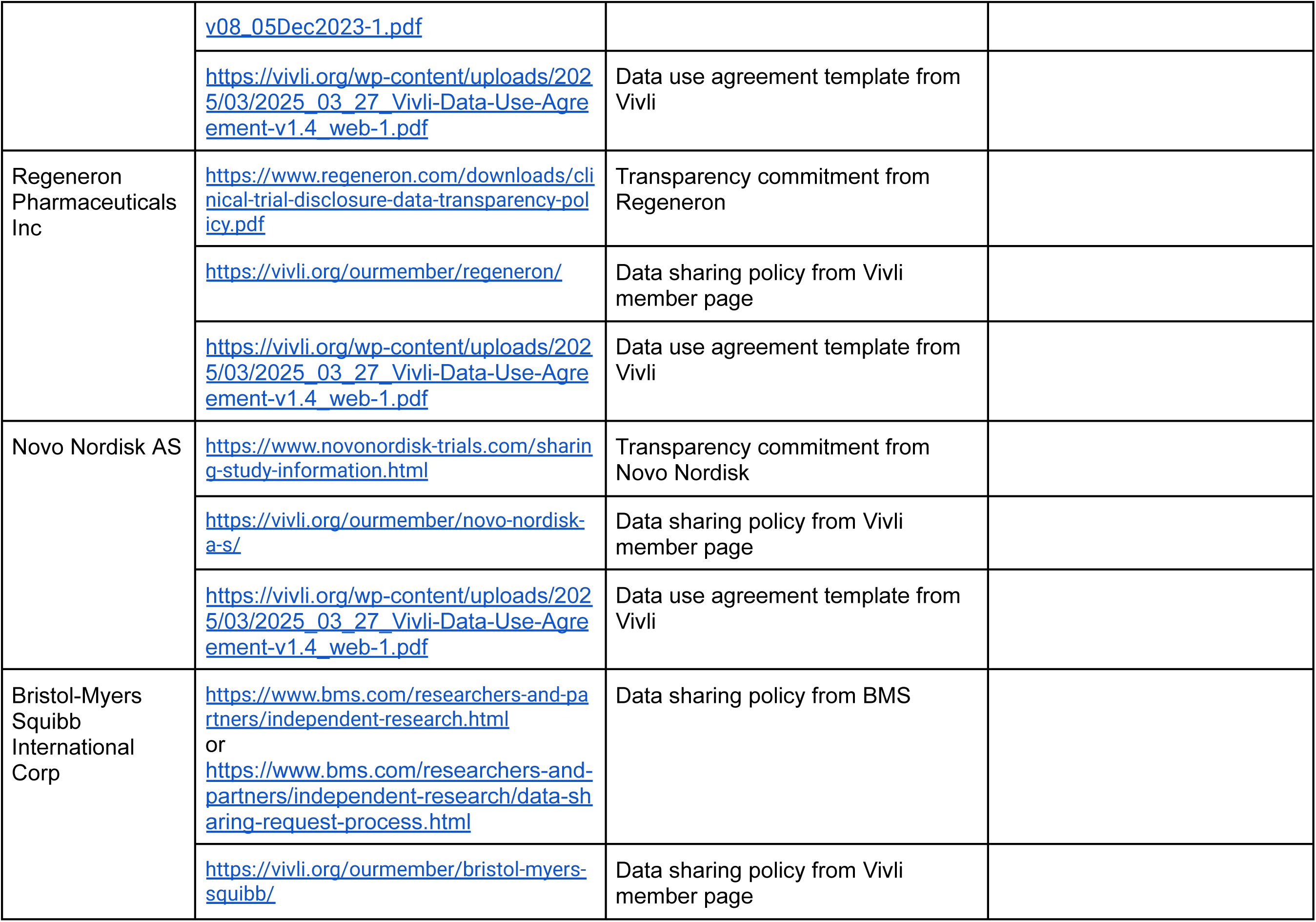
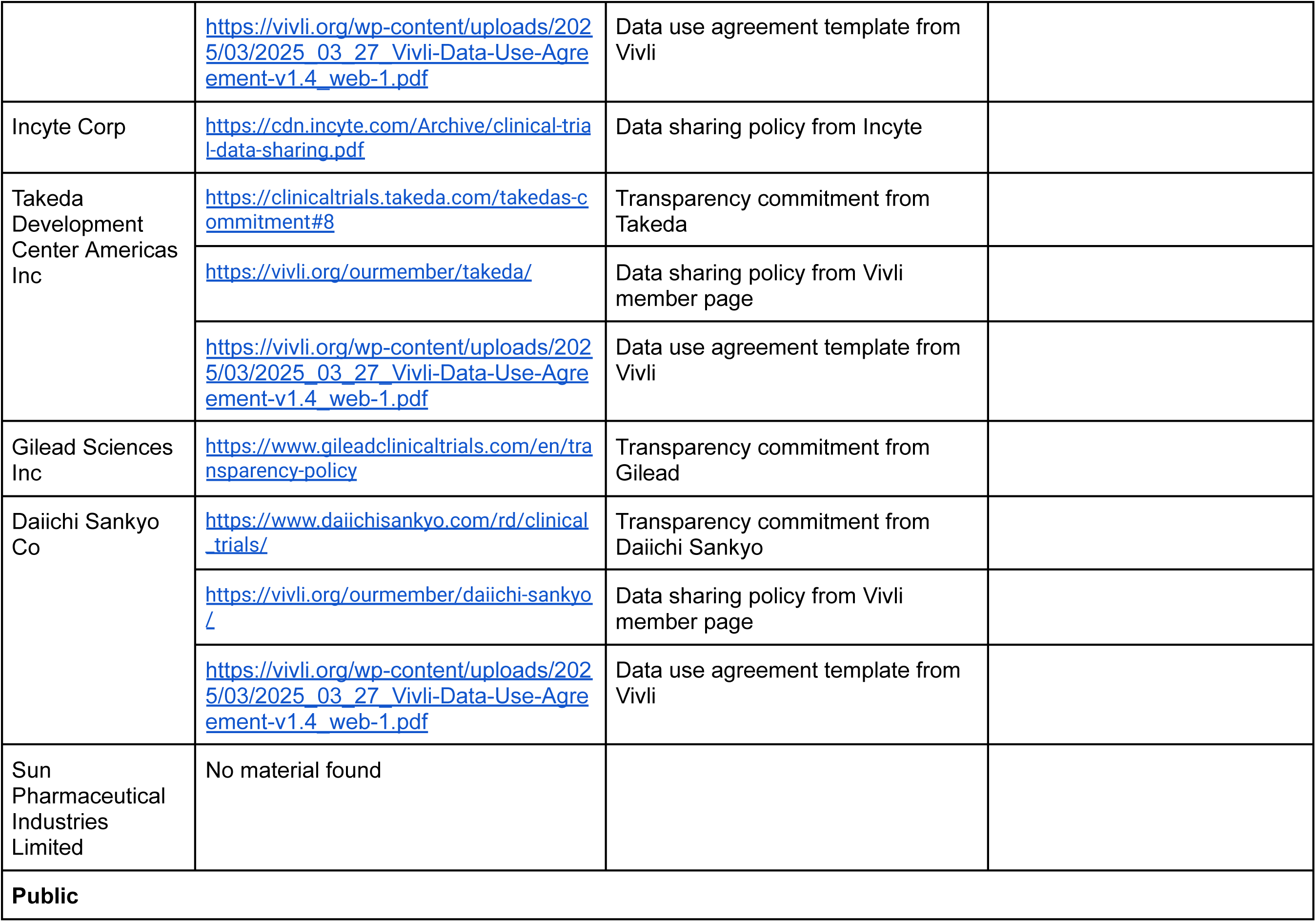
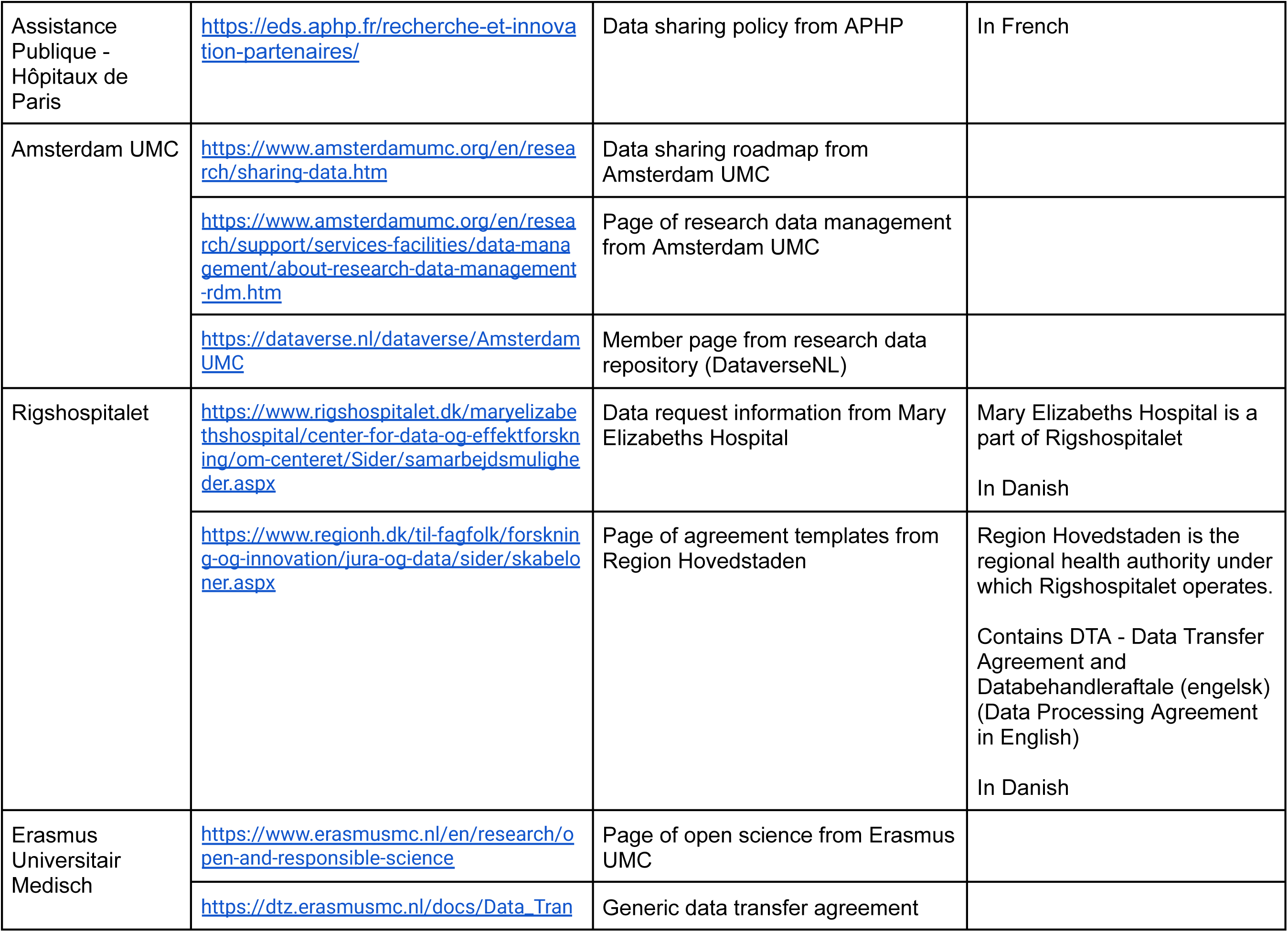
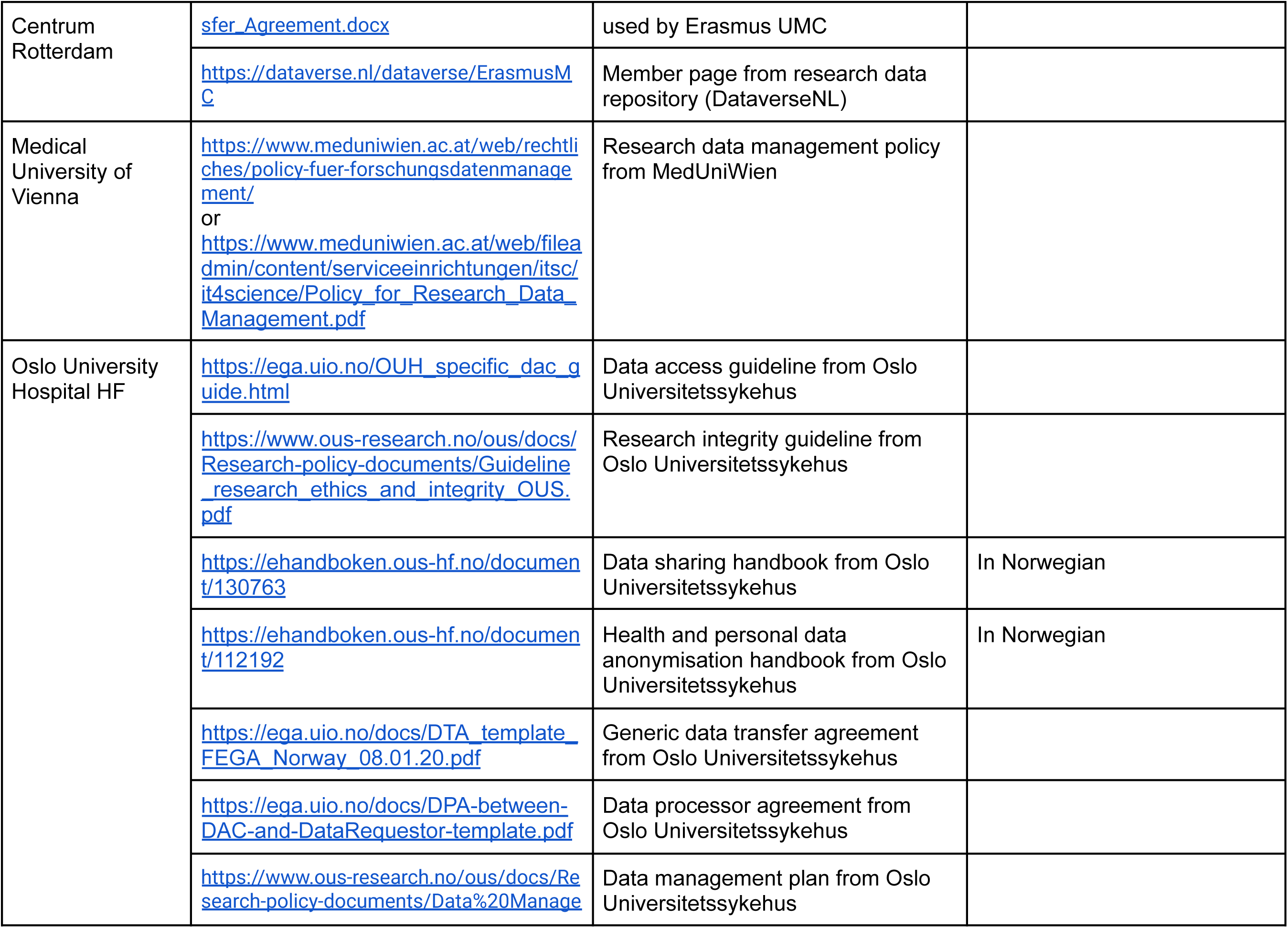
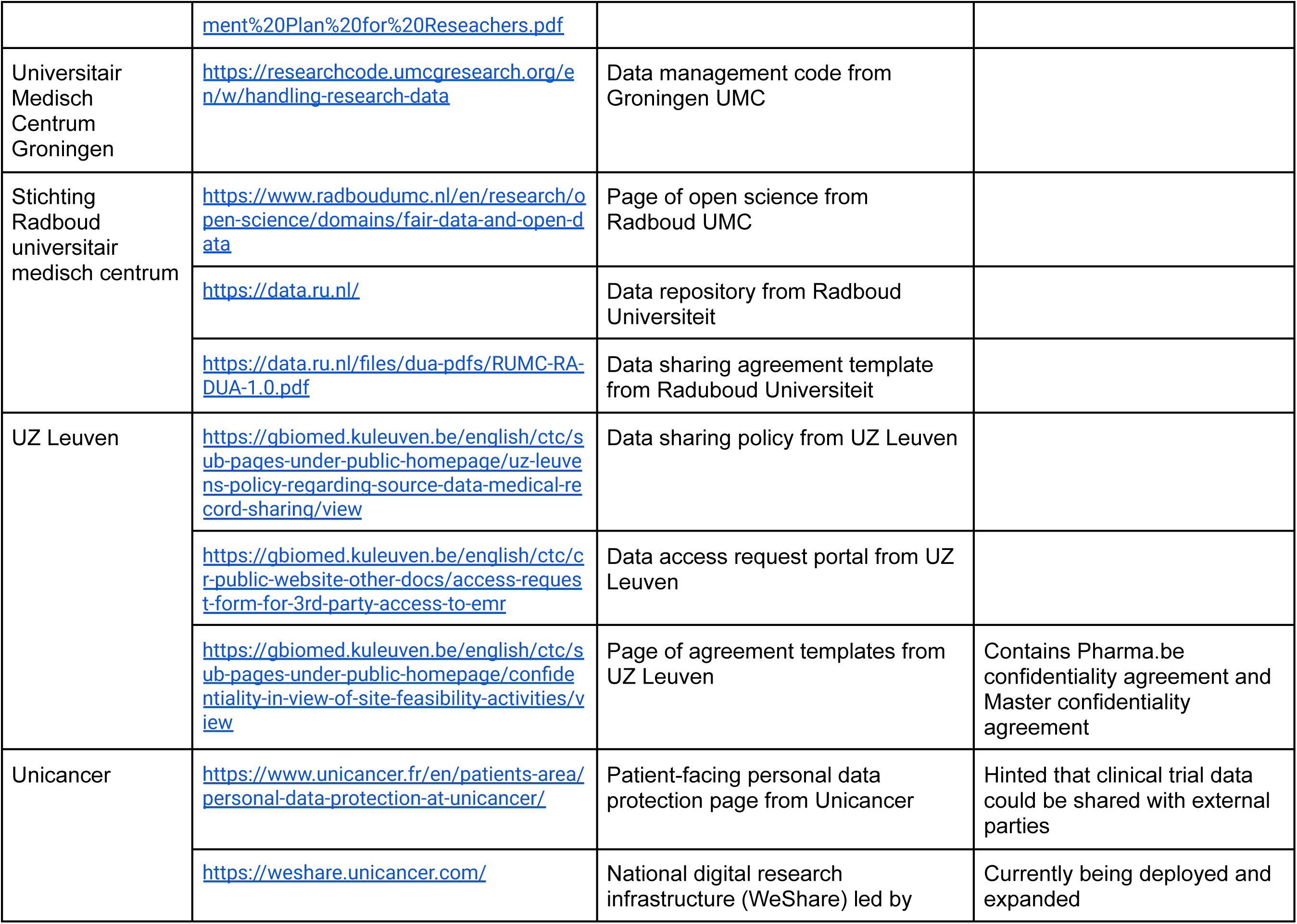
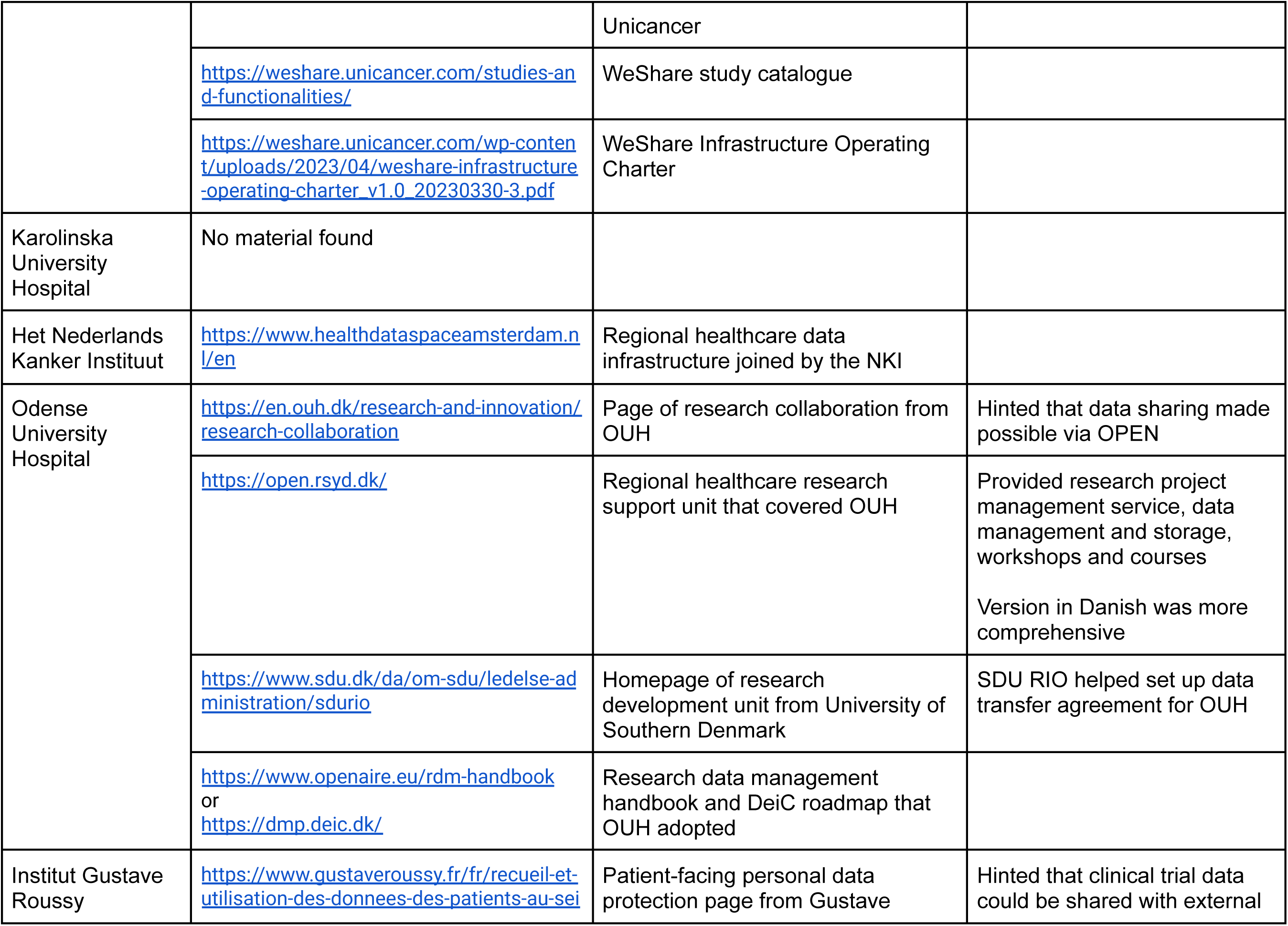
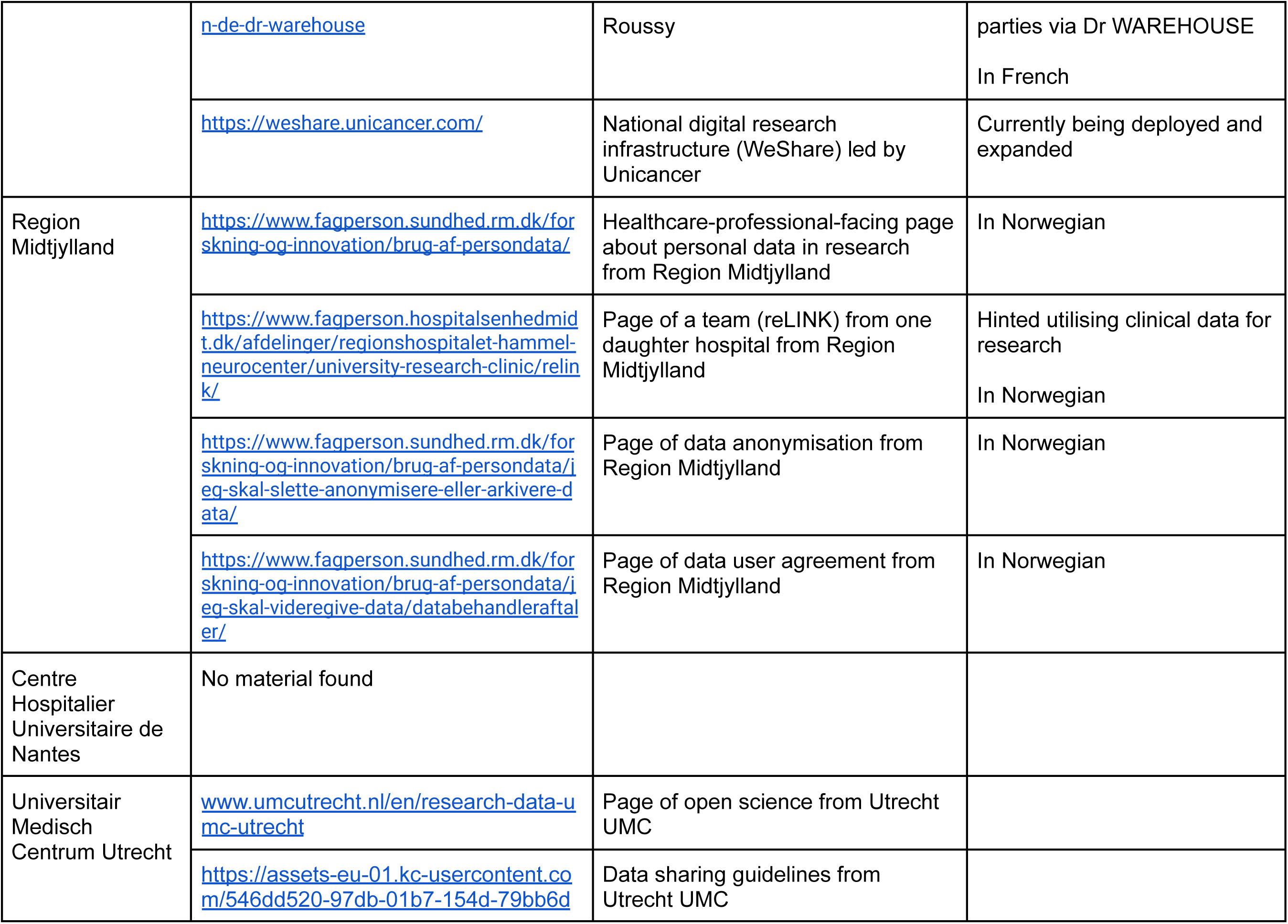
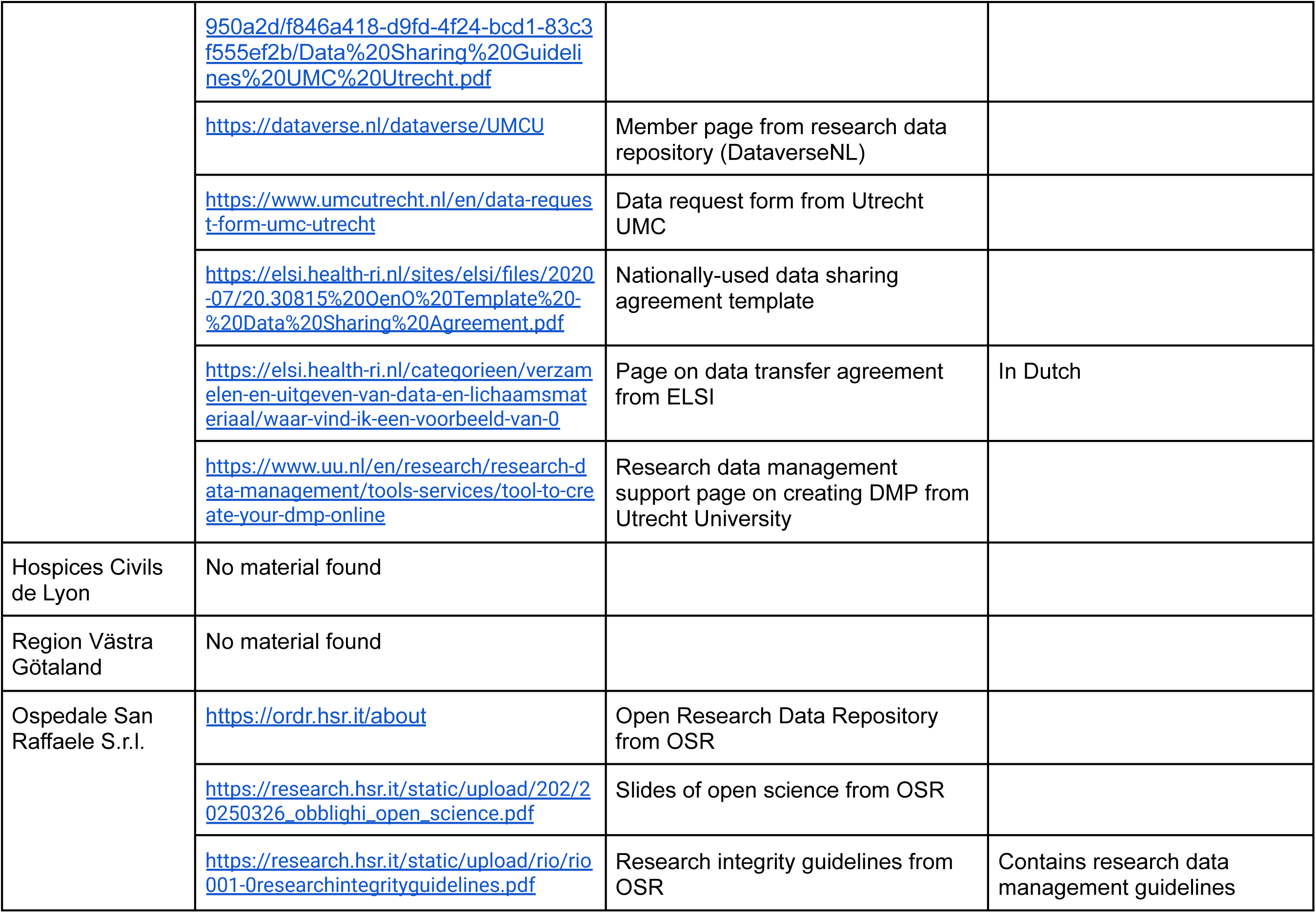
Online material from sponsors assessed for this study.

**Supplementary table 3.**
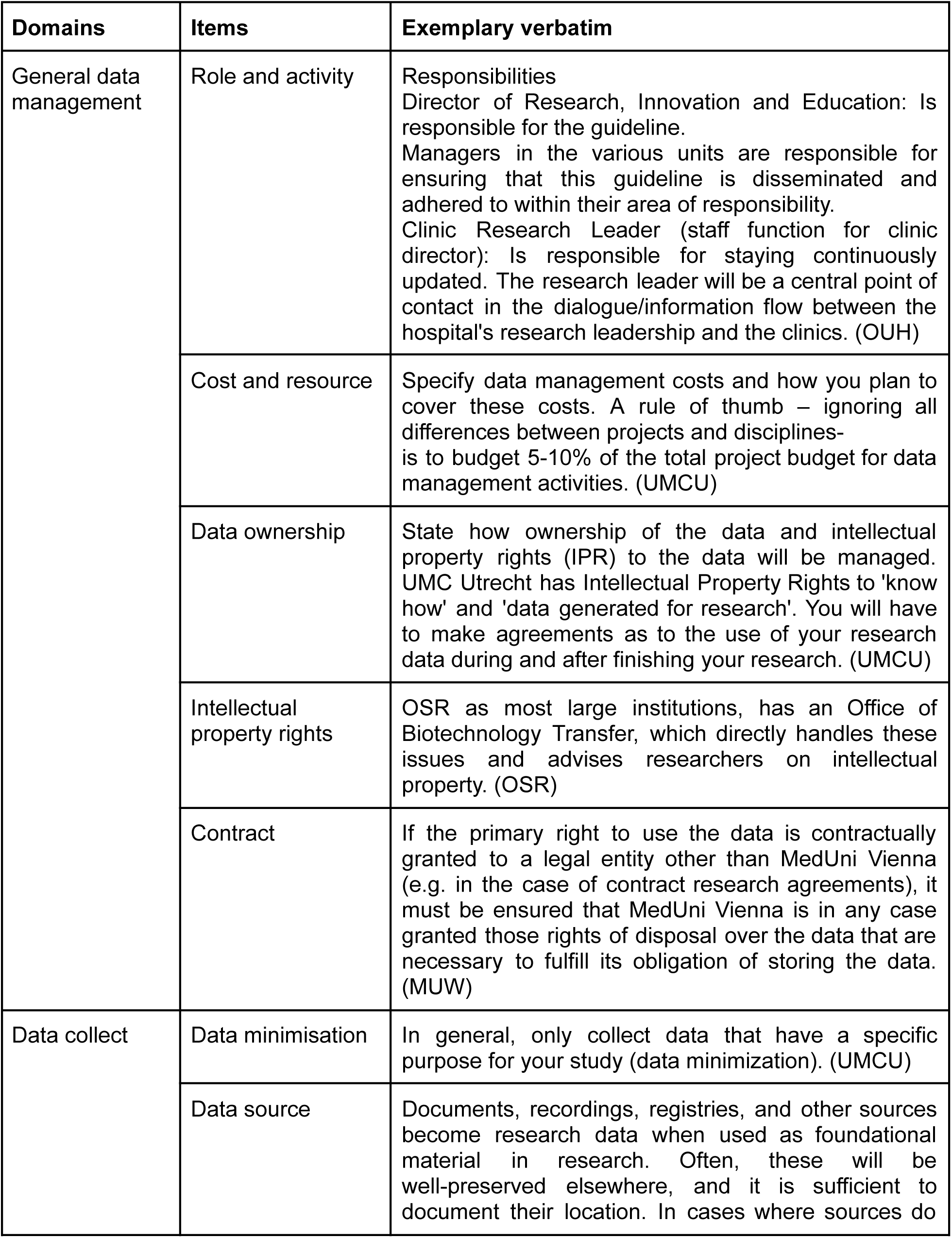
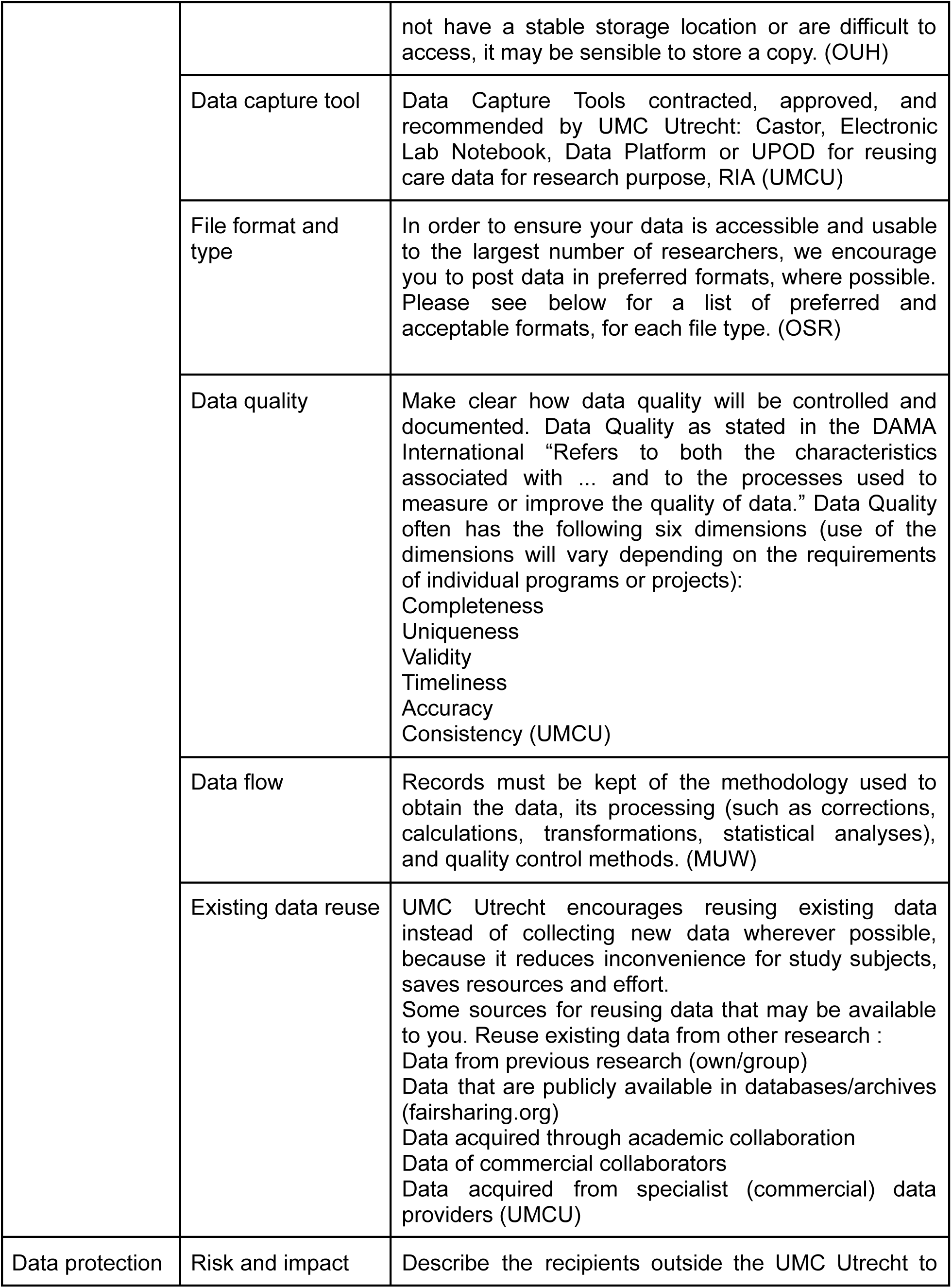
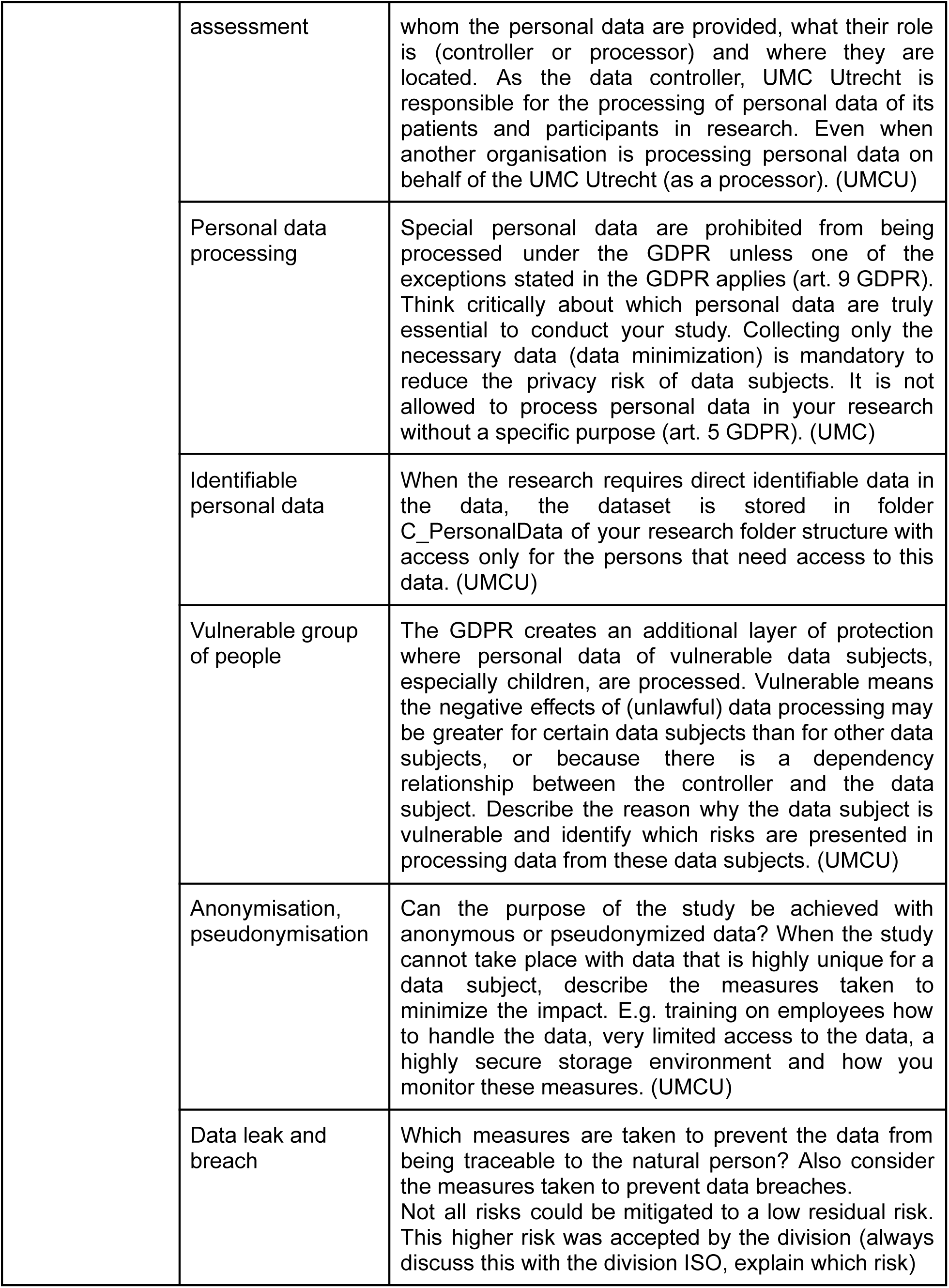
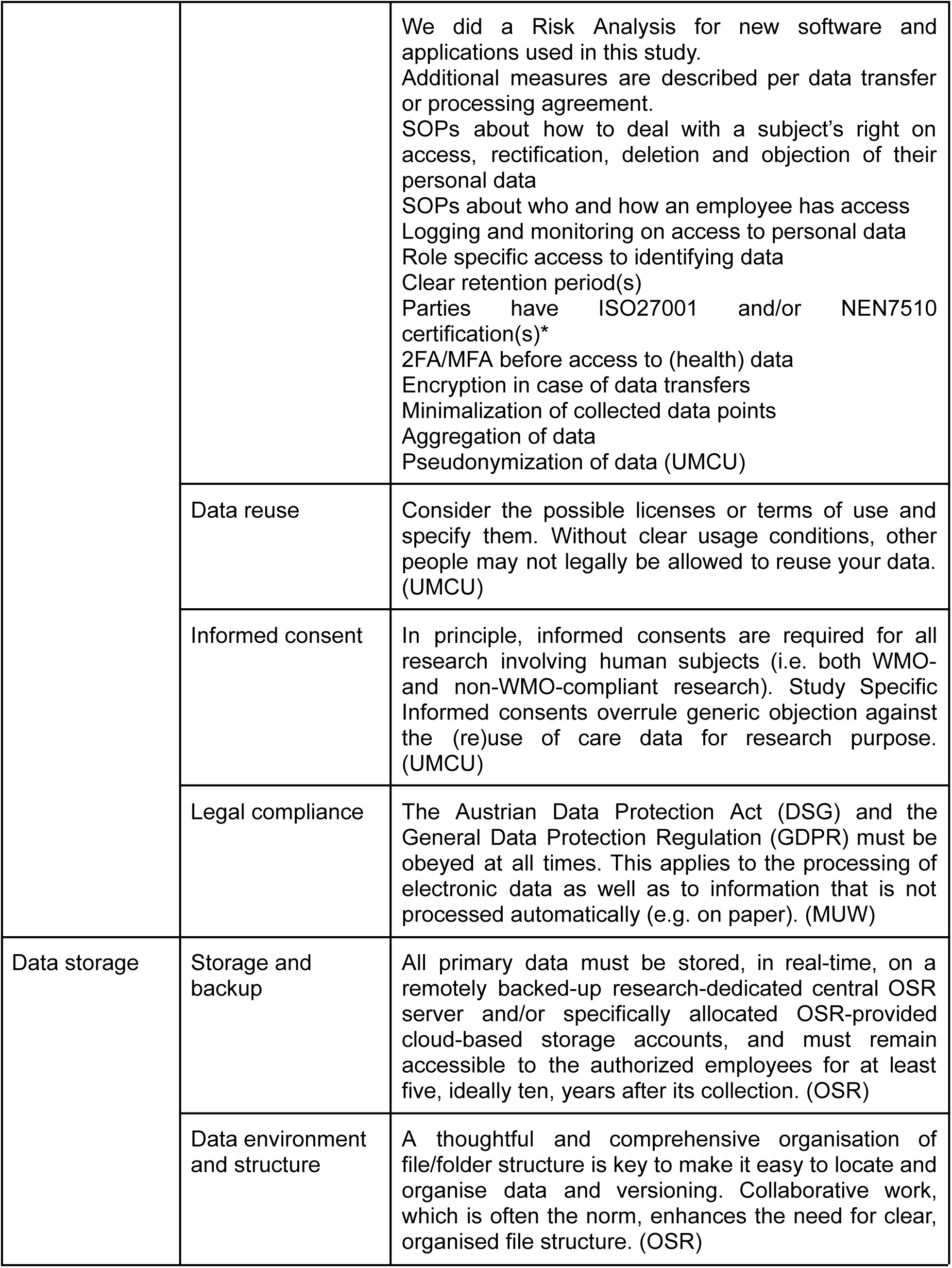
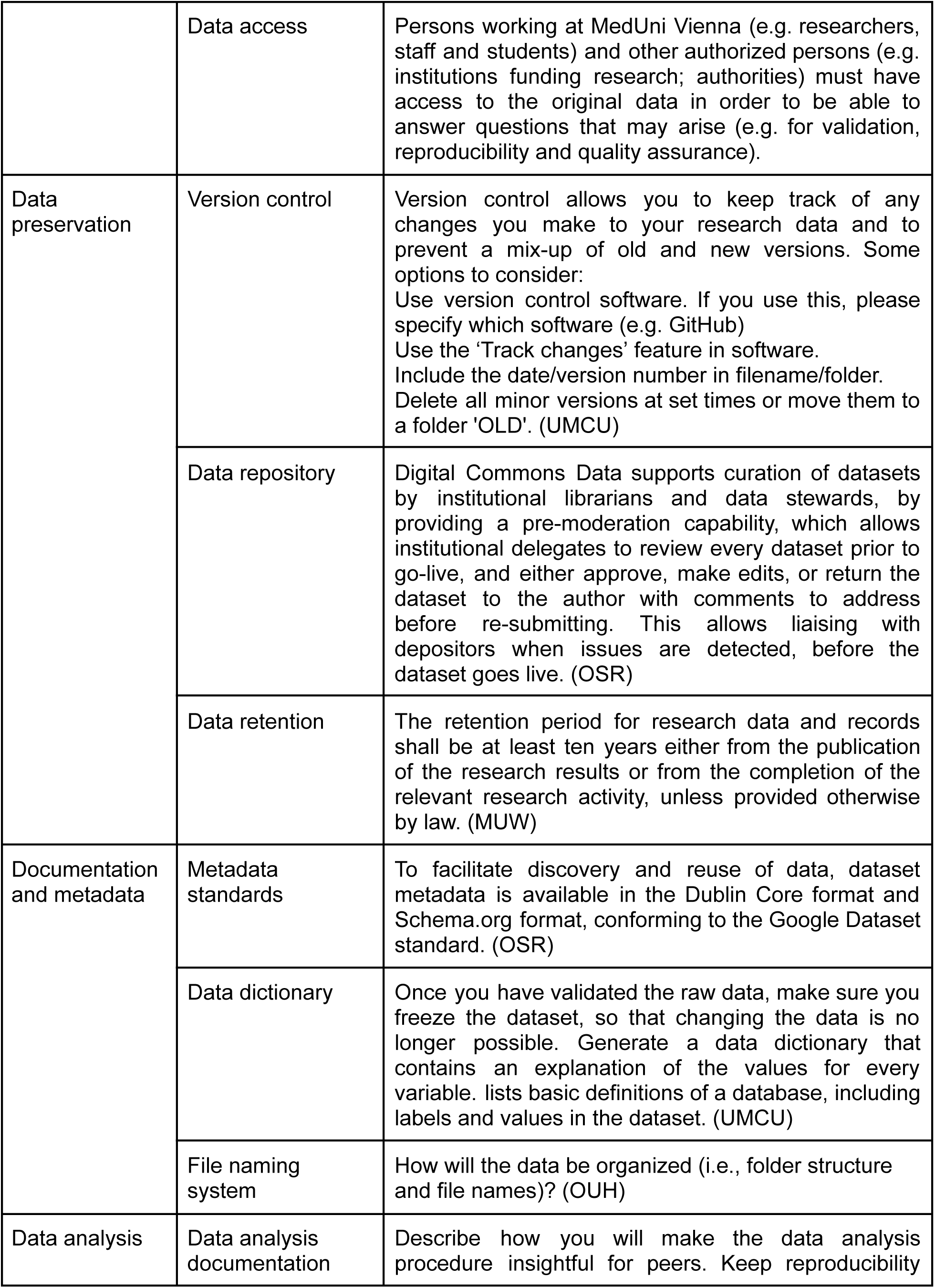
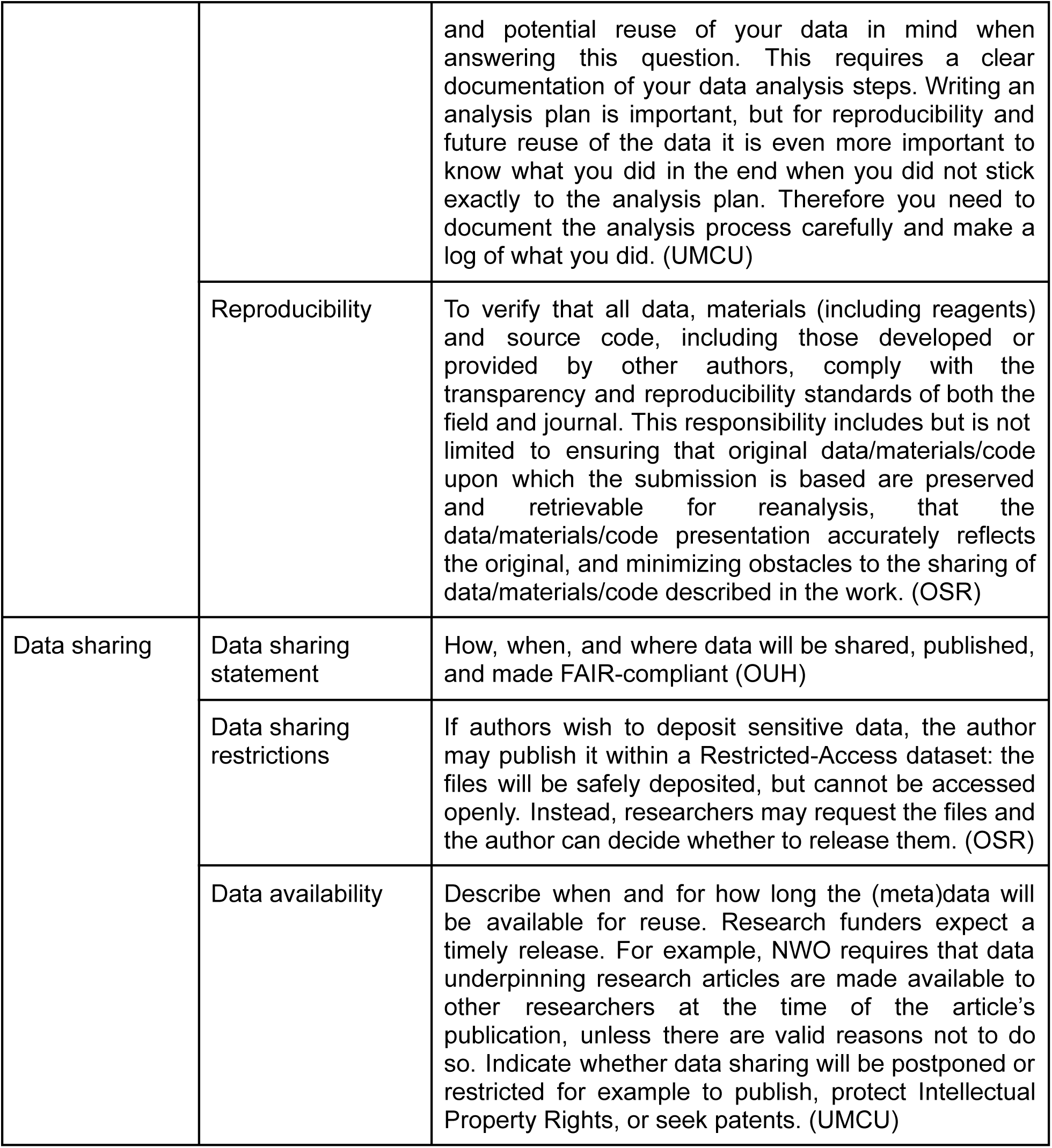
Common items of DMSP found from public sponsors.

**Supplementary table 4.**
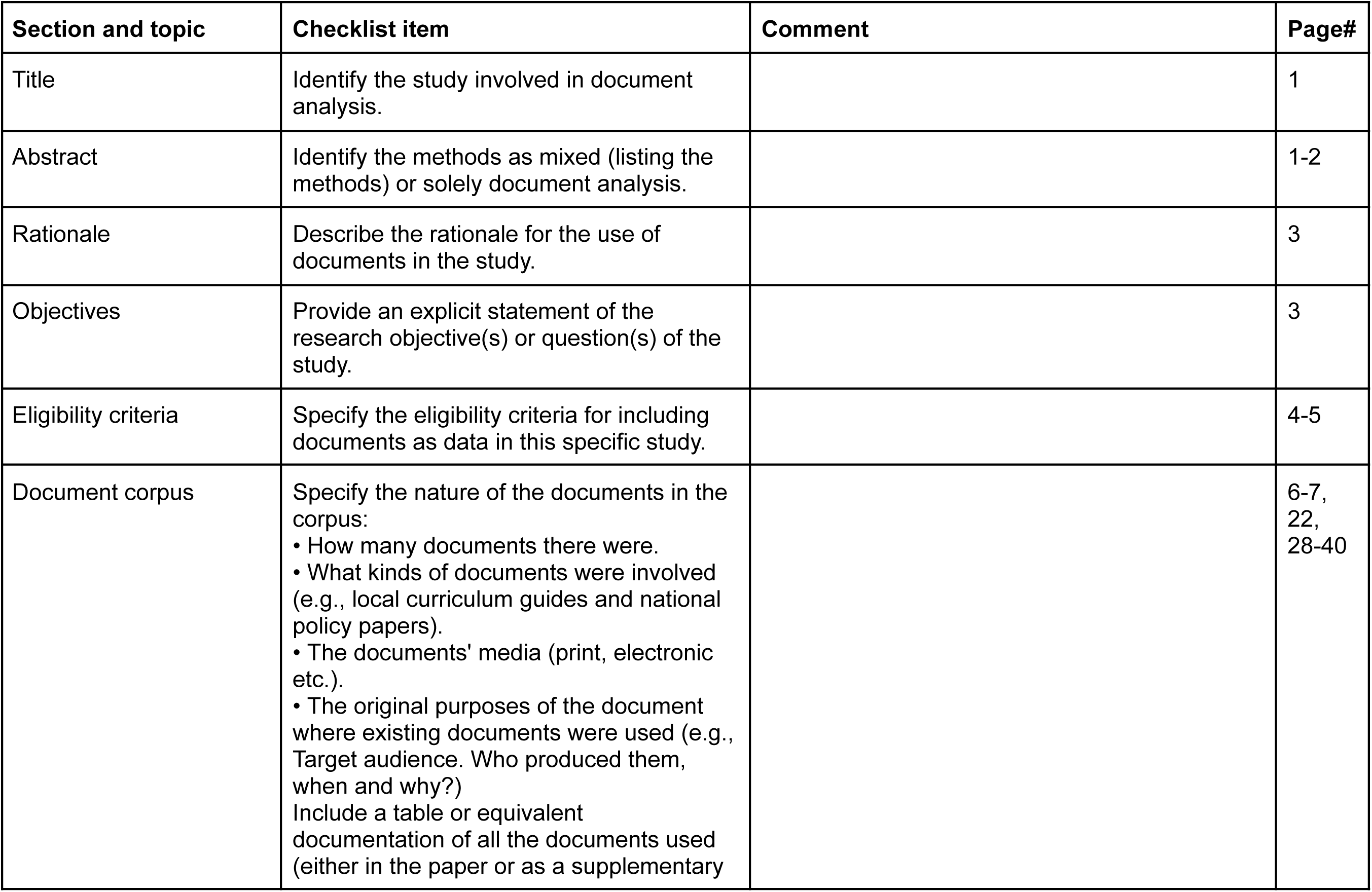
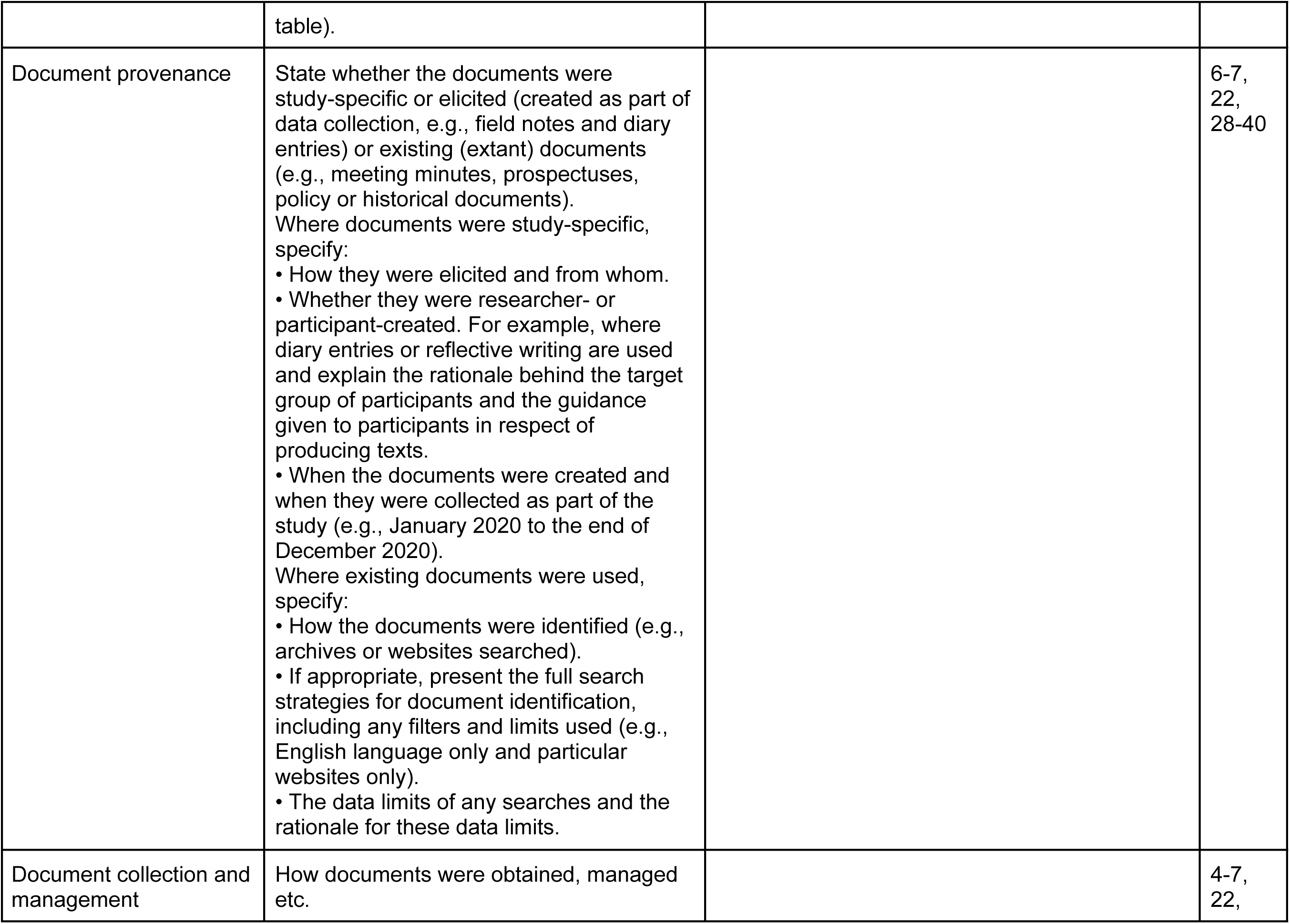
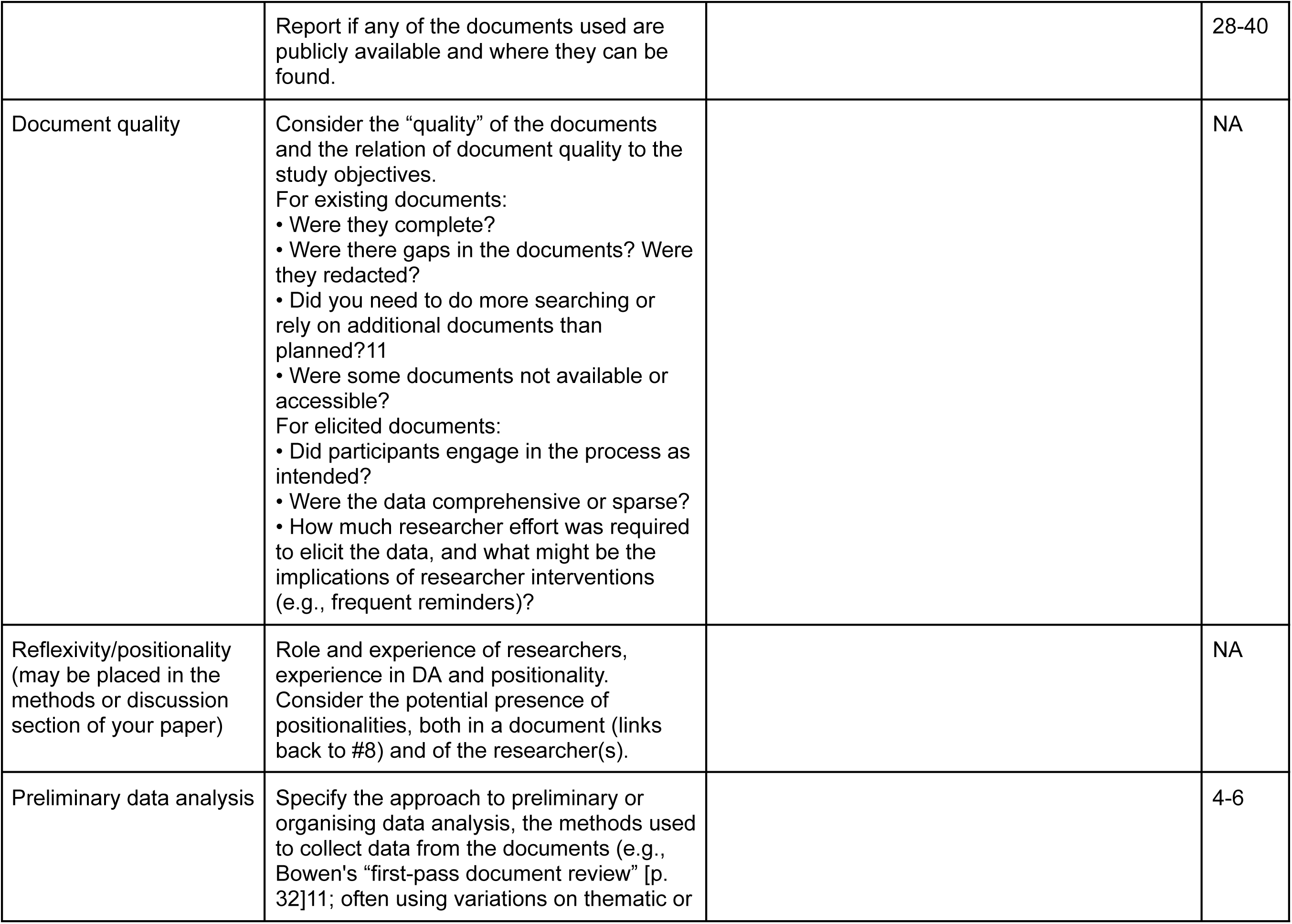
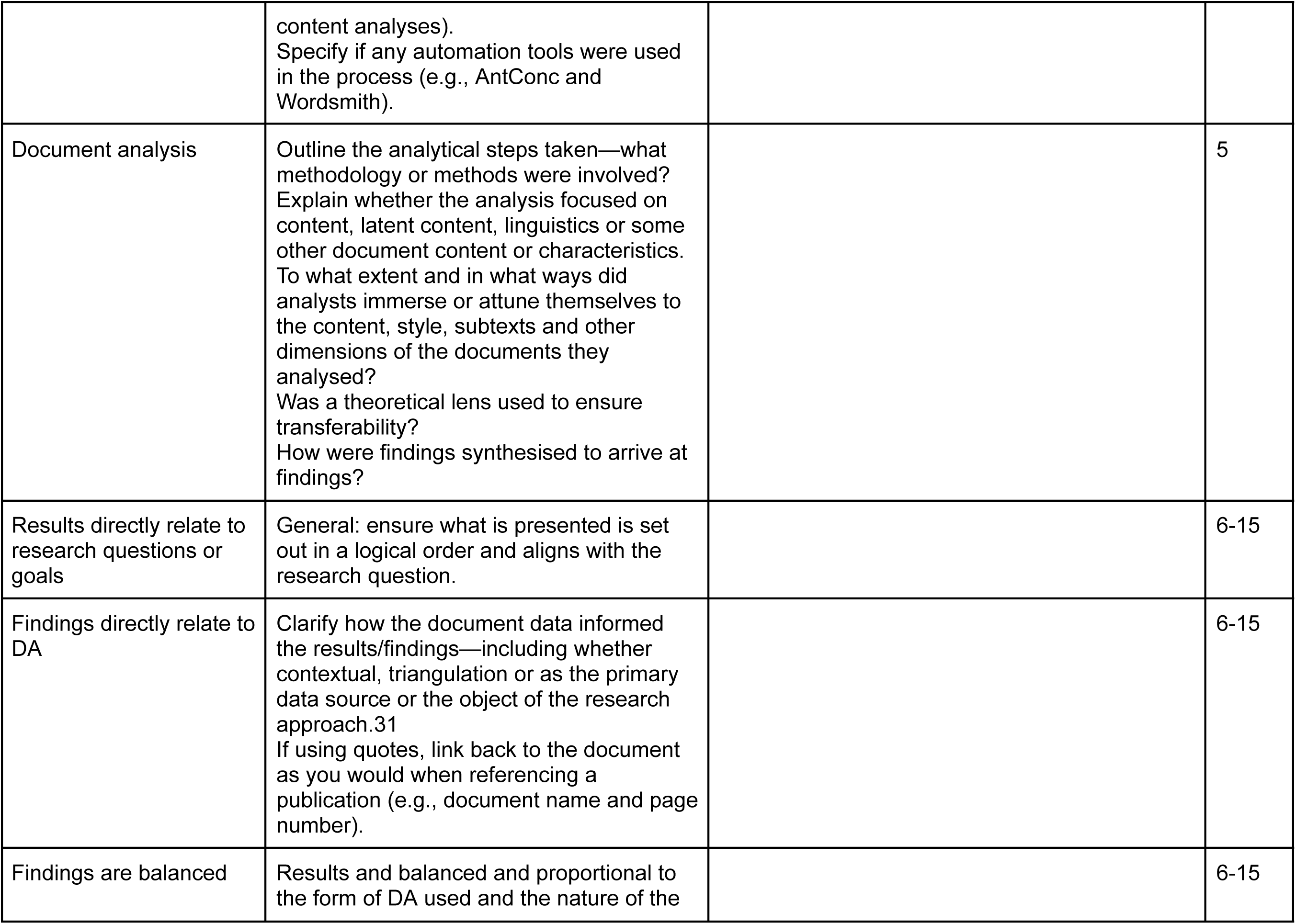
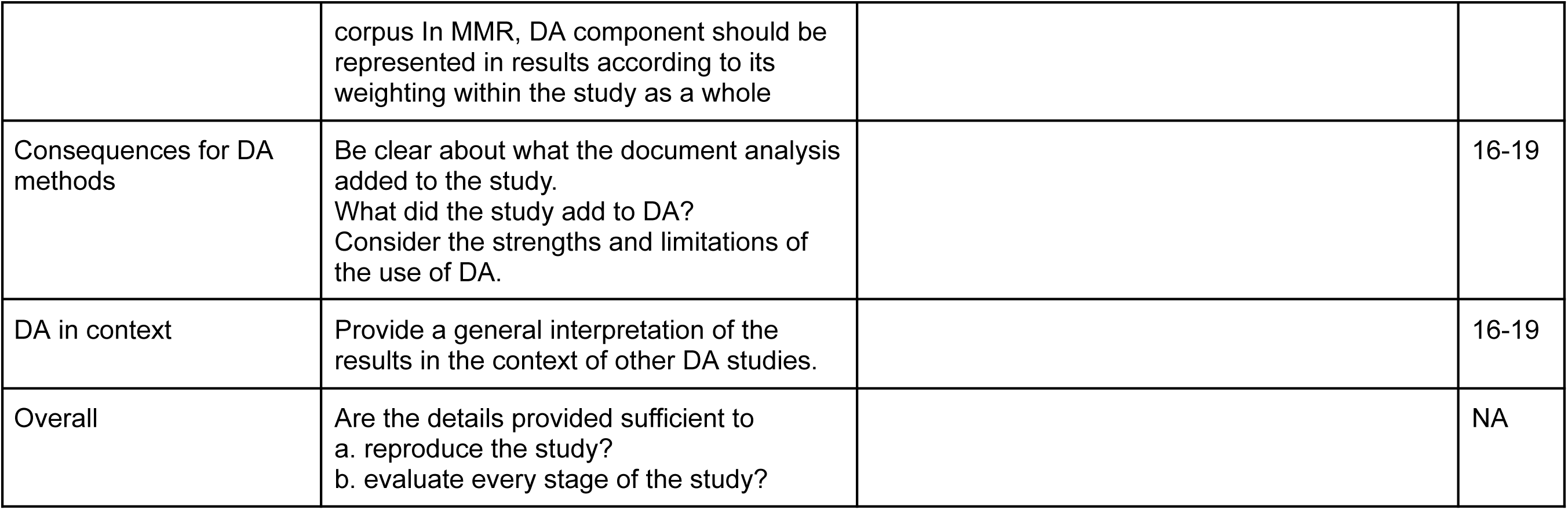
Checklist for Assessment and Reporting of Document Analysis (CARDA)

