## Supplementary material for "Data sharing policies, requirements, and support from public and private clinical trial sponsors: a survey on top sponsors of clinical trials in Europe": Checklist for Assessment and Reporting of Document Analysis (CARDA)

Supplementary table 4 - Checklist for Assessment and Reporting of Document Analysis (CARDA)

| **Section and topic** | **Checklist item** | **Comment** | **Page#** |
|---|---|---|---|
| Title | Identify the study involved in document analysis. | | 1 |
| Abstract | Identify the methods as mixed (listing the methods) or solely document analysis. | | 1 |
| Rationale | Describe the rationale for the use of documents in the study. | | 3 |
| Objectives | Provide an explicit statement of the research objective(s) or question(s) of the study. | | 3 |
| Eligibility criteria | Specify the eligibility criteria for including documents as data in this specific study. | | 4-5 |
| Document corpus | Specify the nature of the documents in the corpus:<br>• How many documents there were.<br>• What kinds of documents were involved (e.g., local curriculum guides and national policy papers).<br>• The documents' media (print, electronic etc.).<br>• The original purposes of the document where existing documents were used (e.g., Target audience. Who produced them, when and why?)<br>Include a table or equivalent documentation of all the documents used (either in the paper or as a supplementary | | 6-7, 19-31 |

| | | | |
|---|---|---|---|
| | table). | | |
| Document provenance | State whether the documents were study-specific or elicited (created as part of data collection, e.g., field notes and diary entries) or existing (extant) documents (e.g., meeting minutes, prospectuses, policy or historical documents).<br>Where documents were study-specific, specify:<br>• How they were elicited and from whom.<br>• Whether they were researcher- or participant-created. For example, where diary entries or reflective writing are used and explain the rationale behind the target group of participants and the guidance given to participants in respect of producing texts.<br>• When the documents were created and when they were collected as part of the study (e.g., January 2020 to the end of December 2020).<br>Where existing documents were used, specify:<br>• How the documents were identified (e.g., archives or websites searched).<br>• If appropriate, present the full search strategies for document identification, including any filters and limits used (e.g., English language only and particular websites only).<br>• The data limits of any searches and the rationale for these data limits. | | 6-7, 19-31 |
| Document collection and management | How documents were obtained, managed etc. | | 4-7, 19-31 |

| | | | |
|---|---|---|---|
| | Report if any of the documents used are publicly available and where they can be found. | | |
| Document quality | Consider the "quality" of the documents and the relation of document quality to the study objectives.<br>For existing documents:<br>• Were they complete?<br>• Were there gaps in the documents? Were they redacted?<br>• Did you need to do more searching or rely on additional documents than planned?11<br>• Were some documents not available or accessible?<br>For elicited documents:<br>• Did participants engage in the process as intended?<br>• Were the data comprehensive or sparse?<br>• How much researcher effort was required to elicit the data, and what might be the implications of researcher interventions (e.g., frequent reminders)? | | NA |
| Reflexivity/positionality (may be placed in the methods or discussion section of your paper) | Role and experience of researchers, experience in DA and positionality. Consider the potential presence of positionalities, both in a document (links back to #8) and of the researcher(s). | | NA |
| Preliminary data analysis | Specify the approach to preliminary or organising data analysis, the methods used to collect data from the documents (e.g., Bowen's "first-pass document review" [p. 32]11; often using variations on thematic or | | 5 |

| | | | |
|---|---|---|---|
| | content analyses).<br>Specify if any automation tools were used in the process (e.g., AntConc and Wordsmith). | | |
| Document analysis | Outline the analytical steps taken—what methodology or methods were involved?<br>Explain whether the analysis focused on content, latent content, linguistics or some other document content or characteristics.<br>To what extent and in what ways did analysts immerse or attune themselves to the content, style, subtexts and other dimensions of the documents they analysed?<br>Was a theoretical lens used to ensure transferability?<br>How were findings synthesised to arrive at findings? | | 5 |
| Results directly relate to research questions or goals | General: ensure what is presented is set out in a logical order and aligns with the research question. | | 6-12 |
| Findings directly relate to DA | Clarify how the document data informed the results/findings—including whether contextual, triangulation or as the primary data source or the object of the research approach.31<br>If using quotes, link back to the document as you would when referencing a publication (e.g., document name and page number). | | 6-12 |
| Findings are balanced | Results and balanced and proportional to the form of DA used and the nature of the | | 6-12 |

| | | | |
|---|---|---|---|
| | corpus In MMR, DA component should be represented in results according to its weighting within the study as a whole | | |
| Consequences for DA methods | Be clear about what the document analysis added to the study.<br>What did the study add to DA?<br>Consider the strengths and limitations of the use of DA. | | 12-15 |
| DA in context | Provide a general interpretation of the results in the context of other DA studies. | | 12-15 |
| Overall | Are the details provided sufficient to<br>a. reproduce the study?<br>b. evaluate every stage of the study? | | NA |
