## Supplementary material for "Data sharing policies, requirements, and support from public and private clinical trial sponsors: a survey on top sponsors of clinical trials in Europe": Common items of Data Management and Sharing Plan found from public sponsors

Supplementary table 3 - Common items of DMSP found from public sponsors

| Domains | Items | Exemplary verbatim |
|---|---|---|
| General data management | Role and activity | Responsibilities<br>Director of Research, Innovation and Education: Is responsible for the guideline.<br>Managers in the various units are responsible for ensuring that this guideline is disseminated and adhered to within their area of responsibility.<br>Clinic Research Leader (staff function for clinic director): Is responsible for staying continuously updated. The research leader will be a central point of contact in the dialogue/information flow between the hospital's research leadership and the clinics. (OUH) |
| | Cost and resource | Specify data management costs and how you plan to cover these costs. A rule of thumb – ignoring all differences between projects and disciplines-<br>is to budget 5-10% of the total project budget for data management activities. (UMCU) |
| | Data ownership | State how ownership of the data and intellectual property rights (IPR) to the data will be managed. UMC Utrecht has Intellectual Property Rights to 'know how' and 'data generated for research'. You will have to make agreements as to the use of your research data during and after finishing your research. (UMCU) |
| | Intellectual property rights | OSR as most large institutions, has an Office of Biotechnology Transfer, which directly handles these issues and advises researchers on intellectual property. (OSR) |
| | Contract | If the primary right to use the data is contractually granted to a legal entity other than MedUni Vienna (e.g. in the case of contract research agreements), it must be ensured that MedUni Vienna is in any case granted those rights of disposal over the data that are necessary to fulfill its obligation of storing the data. (MUW) |
| Data collect | Data minimisation | In general, only collect data that have a specific purpose for your study (data minimization). (UMCU) |
| | Data source | Documents, recordings, registries, and other sources become research data when used as foundational material in research. Often, these will be well-preserved elsewhere, and it is sufficient to document their location. In cases where sources do |

| | | |
|---|---|---|
| | | not have a stable storage location or are difficult to access, it may be sensible to store a copy. (OUH) |
| | Data capture tool | Data Capture Tools contracted, approved, and recommended by UMC Utrecht: Castor, Electronic Lab Notebook, Data Platform or UPOD for reusing care data for research purpose, RIA (UMCU) |
| | File format and type | In order to ensure your data is accessible and usable to the largest number of researchers, we encourage you to post data in preferred formats, where possible. Please see below for a list of preferred and acceptable formats, for each file type. (OSR) |
| | Data quality | Make clear how data quality will be controlled and documented. Data Quality as stated in the DAMA International "Refers to both the characteristics associated with ... and to the processes used to measure or improve the quality of data." Data Quality often has the following six dimensions (use of the dimensions will vary depending on the requirements of individual programs or projects):<br>Completeness<br>Uniqueness<br>Validity<br>Timeliness<br>Accuracy<br>Consistency (UMCU) |
| | Data flow | Records must be kept of the methodology used to obtain the data, its processing (such as corrections, calculations, transformations, statistical analyses), and quality control methods. (MUW) |
| | Existing data reuse | UMC Utrecht encourages reusing existing data instead of collecting new data wherever possible, because it reduces inconvenience for study subjects, saves resources and effort.<br>Some sources for reusing data that may be available to you. Reuse existing data from other research :<br>Data from previous research (own/group)<br>Data that are publicly available in databases/archives (fairsharing.org)<br>Data acquired through academic collaboration<br>Data of commercial collaborators<br>Data acquired from specialist (commercial) data providers (UMCU) |
| Data protection | Risk and impact | Describe the recipients outside the UMC Utrecht to |

| | | |
|---|---|---|
| | assessment | whom the personal data are provided, what their role is (controller or processor) and where they are located. As the data controller, UMC Utrecht is responsible for the processing of personal data of its patients and participants in research. Even when another organisation is processing personal data on behalf of the UMC Utrecht (as a processor). (UMCU) |
| | Personal data processing | Special personal data are prohibited from being processed under the GDPR unless one of the exceptions stated in the GDPR applies (art. 9 GDPR). Think critically about which personal data are truly essential to conduct your study. Collecting only the necessary data (data minimization) is mandatory to reduce the privacy risk of data subjects. It is not allowed to process personal data in your research without a specific purpose (art. 5 GDPR). (UMC) |
| | Identifiable personal data | When the research requires direct identifiable data in the data, the dataset is stored in folder C_PersonalData of your research folder structure with access only for the persons that need access to this data. (UMCU) |
| | Vulnerable group of people | The GDPR creates an additional layer of protection where personal data of vulnerable data subjects, especially children, are processed. Vulnerable means the negative effects of (unlawful) data processing may be greater for certain data subjects than for other data subjects, or because there is a dependency relationship between the controller and the data subject. Describe the reason why the data subject is vulnerable and identify which risks are presented in processing data from these data subjects. (UMCU) |
| | Anonymisation, pseudonymisation | Can the purpose of the study be achieved with anonymous or pseudonymized data? When the study cannot take place with data that is highly unique for a data subject, describe the measures taken to minimize the impact. E.g. training on employees how to handle the data, very limited access to the data, a highly secure storage environment and how you monitor these measures. (UMCU) |
| | Data leak and breach | Which measures are taken to prevent the data from being traceable to the natural person? Also consider the measures taken to prevent data breaches.<br>Not all risks could be mitigated to a low residual risk. This higher risk was accepted by the division (always discuss this with the division ISO, explain which risk) |

| | | |
|---|---|---|
| | | We did a Risk Analysis for new software and applications used in this study.<br>Additional measures are described per data transfer or processing agreement.<br>SOPs about how to deal with a subject's right on access, rectification, deletion and objection of their personal data<br>SOPs about who and how an employee has access<br>Logging and monitoring on access to personal data<br>Role specific access to identifying data<br>Clear retention period(s)<br>Parties have ISO27001 and/or NEN7510 certification(s)*<br>2FA/MFA before access to (health) data<br>Encryption in case of data transfers<br>Minimalization of collected data points<br>Aggregation of data<br>Pseudonymization of data (UMCU) |
| | Data reuse | Consider the possible licenses or terms of use and specify them. Without clear usage conditions, other people may not legally be allowed to reuse your data. (UMCU) |
| | Informed consent | In principle, informed consents are required for all research involving human subjects (i.e. both WMO- and non-WMO-compliant research). Study Specific Informed consents overrule generic objection against the (re)use of care data for research purpose. (UMCU) |
| | Legal compliance | The Austrian Data Protection Act (DSG) and the General Data Protection Regulation (GDPR) must be obeyed at all times. This applies to the processing of electronic data as well as to information that is not processed automatically (e.g. on paper). (MUW) |
| Data storage | Storage and backup | All primary data must be stored, in real-time, on a remotely backed-up research-dedicated central OSR server and/or specifically allocated OSR-provided cloud-based storage accounts, and must remain accessible to the authorized employees for at least five, ideally ten, years after its collection. (OSR) |
| | Data environment and structure | A thoughtful and comprehensive organisation of file/folder structure is key to make it easy to locate and organise data and versioning. Collaborative work, which is often the norm, enhances the need for clear, organised file structure. (OSR) |

| | | |
|---|---|---|
| | Data access | Persons working at MedUni Vienna (e.g. researchers, staff and students) and other authorized persons (e.g. institutions funding research; authorities) must have access to the original data in order to be able to answer questions that may arise (e.g. for validation, reproducibility and quality assurance). |
| Data preservation | Version control | Version control allows you to keep track of any changes you make to your research data and to prevent a mix-up of old and new versions. Some options to consider:<br>Use version control software. If you use this, please specify which software (e.g. GitHub)<br>Use the 'Track changes' feature in software.<br>Include the date/version number in filename/folder.<br>Delete all minor versions at set times or move them to a folder 'OLD'. (UMCU) |
| | Data repository | Digital Commons Data supports curation of datasets by institutional librarians and data stewards, by providing a pre-moderation capability, which allows institutional delegates to review every dataset prior to go-live, and either approve, make edits, or return the dataset to the author with comments to address before re-submitting. This allows liaising with depositors when issues are detected, before the dataset goes live. (OSR) |
| | Data retention | The retention period for research data and records shall be at least ten years either from the publication of the research results or from the completion of the relevant research activity, unless provided otherwise by law. (MUW) |
| Documentation and metadata | Metadata standards | To facilitate discovery and reuse of data, dataset metadata is available in the Dublin Core format and Schema.org format, conforming to the Google Dataset standard. (OSR) |
| | Data dictionary | Once you have validated the raw data, make sure you freeze the dataset, so that changing the data is no longer possible. Generate a data dictionary that contains an explanation of the values for every variable. lists basic definitions of a database, including labels and values in the dataset. (UMCU) |
| | File naming system | How will the data be organized (i.e., folder structure and file names)? (OUH) |
| Data analysis | Data analysis documentation | Describe how you will make the data analysis procedure insightful for peers. Keep reproducibility |

| | | |
|---|---|---|
| | | and potential reuse of your data in mind when answering this question. This requires a clear documentation of your data analysis steps. Writing an analysis plan is important, but for reproducibility and future reuse of the data it is even more important to know what you did in the end when you did not stick exactly to the analysis plan. Therefore you need to document the analysis process carefully and make a log of what you did. (UMCU) |
| | Reproducibility | To verify that all data, materials (including reagents) and source code, including those developed or provided by other authors, comply with the transparency and reproducibility standards of both the field and journal. This responsibility includes but is not limited to ensuring that original data/materials/code upon which the submission is based are preserved and retrievable for reanalysis, that the data/materials/code presentation accurately reflects the original, and minimizing obstacles to the sharing of data/materials/code described in the work. (OSR) |
| Data sharing | Data sharing statement | How, when, and where data will be shared, published, and made FAIR-compliant (OUH) |
| | Data sharing restrictions | If authors wish to deposit sensitive data, the author may publish it within a Restricted-Access dataset: the files will be safely deposited, but cannot be accessed openly. Instead, researchers may request the files and the author can decide whether to release them. (OSR) |
| | Data availability | Describe when and for how long the (meta)data will be available for reuse. Research funders expect a timely release. For example, NWO requires that data underpinning research articles are made available to other researchers at the time of the article's publication, unless there are valid reasons not to do so. Indicate whether data sharing will be postponed or restricted for example to publish, protect Intellectual Property Rights, or seek patents. (UMCU) |
