## Supplementary material for "Data sharing policies, requirements, and support from public and private clinical trial sponsors: a survey on top sponsors of clinical trials in Europe": Online material from sponsors assessed for this study

Supplementary table 2 - Online material from sponsors assessed for this study

| Sponsors | Website | Content | Remark |
|---|---|---|---|
| **Private** | | | |
| Merck Sharp & Dohme LLC | https://trialstransparency.msdclinicaltrials.com/pdf/ProcedureAccessClinicalTrialData.pdf | Data sharing policy from MSD | |
| | https://externaldatasharing-msd.com/ or https://www.msd-trialdatasharing-dev.com/ | Data request site from MSD | |
| | https://vivli.org/ourmember/merck-sharp-dohme-llc-rahway-nj-usa/ | Data sharing policy from Vivli member page | MSD required data seekers to submit a request proposal on their MSD platform. Upon approval, data will be uploaded to Vivli for access. |
| | https://vivli.org/wp-content/uploads/2025/03/2025_03_27_Vivli-Data-Use-Agreement-v1.4_web-1.pdf | Data use agreement template from Vivli | |
| AstraZeneca AB | https://www.astrazenecaclinicaltrials.com/our-transparency-commitments/ | Transparency commitments from AstraZeneca site | |
| | https://vivli.org/ourmember/astrazeneca/ | Data sharing policy from Vivli member page | |
| | https://vivli.org/wp-content/uploads/2025/03/2025_03_27_Vivli-Data-Use-Agreement-v1.4_web-1.pdf | Data use agreement template from Vivli | |
| Novartis Pharma AG | https://www.novartis.com/clinicaltrials/transparency | Transparency commitments from Novartis site | |

| | | |
|---|---|---|
| | https://www.clinicalstudydatarequest.com/Study-Sponsors/Study-Sponsors-Novartis.aspx | Data sharing policy from CSDR member page |
| | https://vivli.org/ourmember/novartis-pharma-ag/ | Data sharing policy from Vivli member page |
| | https://www.clinicalstudydatarequest.com/Documents/Novartis-Global-Clinical%20-Data-Anonymization-Standards%20(v4).pdf | Data anonymisation standards from Novartis |
| | https://www.clinicalstudydatarequest.com/Help/Help-Data-Sharing-Agreement.aspx | Data sharing agreement explanation page from CSDR |
| | https://www.clinicalstudydatarequest.com/Documents/CSDR%20Single%20and%20Multi-sponsor%20DSA_V9.pdf | Data sharing agreement template from CSDR |
| | https://vivli.org/wp-content/uploads/2025/03/2025_03_27_Vivli-Data-Use-Agreement-v1.4_web-1.pdf | Data use agreement template from Vivli |
| F. Hoffmann-La Roche | https://www.roche.com/innovation/process/clinical-trials/data-sharing | Transparency commitment from Roche site |
| | https://vivli.org/ourmember/roche/ | Data sharing policy from Vivli member page |
| | https://vivli.org/wp-content/uploads/2023/10/Anonymization-of-clinical-trial-data-for-sharing-via-Vivli_Final-20APR2021.pdf | Data anonymisation framework from Roche |
| | https://vivli.org/wp-content/uploads/2025/03/2025_03_27_Vivli-Data-Use-Agreement-v1.4_web-1.pdf | Data use agreement template from Vivli |

| | | | |
|---|---|---|---|
| Janssen-Cilag International | https://www.jnj.com/policies-reports/our-position-on-clinical-trial-data-transparency | Transparency commitment from Johnson & Johnson | |
| | https://vivli.org/ourmember/johnson-johnson/ | Data sharing policy from Vivli member page | Vivli page served as an advertisement and pointed data seekers to YODA portal |
| | https://yoda.yale.edu/jj-available-data/ | Data sharing policy from YODA member page | |
| | https://yoda.yale.edu/wp-content/uploads/2024/02/YODA-Project-Janssen-DUA-January-2024.pdf or https://yoda.yale.edu/wp-content/uploads/2024/02/YODA-Project-Janssen-Medical-Device-DUA-January-2024.pdf | Data use agreement from YODA with Johnson & Johnson | |
| Boehringer Ingelheim International GmbH | https://www.mystudywindow.com/msw/datasharing | Data sharing policy from Boehringer | |
| | https://www.mystudywindow.com/msw/commitment | Transparency commitment from Boehringer | |
| | https://vivli.org/ourmember/boehringer-ingelheim/ | Data sharing policy from Vivli member page | |
| | https://vivli.org/wp-content/uploads/2025/03/2025_03_11-Boehringer-Ingelheim-Data-Protection-Procedures.pdf | Data protection procedures from Boehringer | |
| | https://vivli.org/wp-content/uploads/2025/03/2025_03_27_Vivli-Data-Use-Agreement-v1.4_web-1.pdf | Data use agreement template from Vivli | |
| Pfizer Inc | https://www.pfizer.com/science/clinical-trials/trial-data-and-results/data-reques | Data sharing policy from Pfizer | |

| | | |
|---|---|---|
| | ts | |
| | https://vivli.org/ourmember/pfizer/ | Data sharing policy from Vivli member page |
| | https://vivli.org/wp-content/uploads/2025/03/2025_03_27_Vivli-Data-Use-Agreement-v1.4_web-1.pdf | Data use agreement template from Vivli |
| AbbVie Deutschland GmbH & Co KG | https://www.abbvieclinicaltrials.com/hcp/data-sharing/ | Data sharing policy from AbbVie |
| | https://vivli.org/ourmember/abbvie/ | Data sharing policy from Vivli member page |
| | https://vivli.org/wp-content/uploads/2025/03/2025_03_27_Vivli-Data-Use-Agreement-v1.4_web-1.pdf | Data use agreement template from Vivli |
| Eli Lilly & Co | https://www.lilly.com/science/clinical-trials | Transparency commitment from Lilly |
| | https://vivli.org/ourmember/lilly/ | Data sharing policy from Vivli member page |
| | https://vivli.org/wp-content/uploads/2025/03/2025_03_27_Vivli-Data-Use-Agreement-v1.4_web-1.pdf | Data use agreement template from Vivli |
| Sanofi-Aventis Recherche & Developpement | https://www.sanofi.com/en/our-science/clinical-trials-and-results/our-data-sharing-commitments | Data sharing policy from Sanofi |
| | https://www.sanofi.com/en/our-company/sustainability/responsible-business-values/bioethics-framework/principles-on-clinical-study-transparency | Transparency commitment from Sanofi |

| | | | |
|---|---|---|---|
| | https://vivli.org/ourmember/sanofi/ | Data sharing policy from Vivli member page | |
| | https://vivli.org/wp-content/uploads/2022/02/Sanofi-Anonymization-standards_October-2025.pdf | Data anonymisation standards from Sanofi | |
| | https://vivli.org/wp-content/uploads/2025/03/2025_03_27_Vivli-Data-Use-Agreement-v1.4_web-1.pdf | Data use agreement template from Vivli | |
| GlaxoSmithKline Research & Development Limited | https://d3l8i7lo48obsd.cloudfront.net/gsk-patient-level-data-sharing-july2025-1-Bgwa1UthxvluYbWYTThw.pdf | Data sharing policy from GSK | Explained that starting in 2023 all new data requests are directed to the Vivli |
| | https://www.gsk-studyregister.com/ | GSK trial registry portal | |
| | https://vivli.org/ourmember/gsk/ | Data sharing policy from Vivli member page | |
| | https://www.clinicalstudydatarequest.com/Study-Sponsors/Study-Sponsors-GSK.aspx | Data sharing policy from CSDR member page | Starting in 2023, all new data requests are directed to the Vivli platform. |
| | https://vivli.org/wp-content/uploads/2025/03/2025_03_27_Vivli-Data-Use-Agreement-v1.4_web-1.pdf | Data use agreement template from Vivli | |
| Bayer AG | https://clinicaltrials.bayer.com/transparency-policy | Transparency commitment from Bayer | |
| | https://vivli.org/ourmember/bayer/ | Data sharing policy from Vivli member page | |
| | https://vivli.org/wp-content/uploads/2023/12/Bayer-Clinical-Trial-Data-Transparency_Data-Anonymization-Process_ | Data anonymisation procedures from Bayer | |

| | | |
|---|---|---|
| | v08_05Dec2023-1.pdf | |
| | https://vivli.org/wp-content/uploads/2025/03/2025_03_27_Vivli-Data-Use-Agreement-v1.4_web-1.pdf | Data use agreement template from Vivli |
| Regeneron Pharmaceuticals Inc | https://www.regeneron.com/downloads/clinical-trial-disclosure-data-transparency-policy.pdf | Transparency commitment from Regeneron |
| | https://vivli.org/ourmember/regeneron/ | Data sharing policy from Vivli member page |
| | https://vivli.org/wp-content/uploads/2025/03/2025_03_27_Vivli-Data-Use-Agreement-v1.4_web-1.pdf | Data use agreement template from Vivli |
| Novo Nordisk AS | https://www.novonordisk-trials.com/sharing-study-information.html | Transparency commitment from Novo Nordisk |
| | https://vivli.org/ourmember/novo-nordisk-a-s/ | Data sharing policy from Vivli member page |
| | https://vivli.org/wp-content/uploads/2025/03/2025_03_27_Vivli-Data-Use-Agreement-v1.4_web-1.pdf | Data use agreement template from Vivli |
| Bristol-Myers Squibb International Corp | https://www.bms.com/researchers-and-partners/independent-research.html or https://www.bms.com/researchers-and-partners/independent-research/data-sharing-request-process.html | Data sharing policy from BMS |
| | https://vivli.org/ourmember/bristol-myers-squibb/ | Data sharing policy from Vivli member page |

| | | |
|---|---|---|
| | https://vivli.org/wp-content/uploads/2025/03/2025_03_27_Vivli-Data-Use-Agreement-v1.4_web-1.pdf | Data use agreement template from Vivli |
| Incyte Corp | https://cdn.incyte.com/Archive/clinical-trial-data-sharing.pdf | Data sharing policy from Incyte |
| Takeda Development Center Americas Inc | https://clinicaltrials.takeda.com/takedas-commitment#8 | Transparency commitment from Takeda |
| | https://vivli.org/ourmember/takeda/ | Data sharing policy from Vivli member page |
| | https://vivli.org/wp-content/uploads/2025/03/2025_03_27_Vivli-Data-Use-Agreement-v1.4_web-1.pdf | Data use agreement template from Vivli |
| Gilead Sciences Inc | https://www.gileadclinicaltrials.com/en/transparency-policy | Transparency commitment from Gilead |
| Daiichi Sankyo Co | https://www.daiichisankyo.com/rd/clinical_trials/ | Transparency commitment from Daiichi Sankyo |
| | https://vivli.org/ourmember/daiichi-sankyo/ | Data sharing policy from Vivli member page |
| | https://vivli.org/wp-content/uploads/2025/03/2025_03_27_Vivli-Data-Use-Agreement-v1.4_web-1.pdf | Data use agreement template from Vivli |
| Sun Pharmaceutical Industries Limited | No material found | |
| **Public** | | |

| | | | |
|---|---|---|---|
| Assistance Publique - Hôpitaux de Paris | https://eds.aphp.fr/recherche-et-innovation-partenaires/ | Data sharing policy from APHP | In French |
| Amsterdam UMC | https://www.amsterdamumc.org/en/research/sharing-data.htm | Data sharing roadmap from Amsterdam UMC | |
| | https://www.amsterdamumc.org/en/research/support/services-facilities/data-management/about-research-data-management-rdm.htm | Page of research data management from Amsterdam UMC | |
| | https://dataverse.nl/dataverse/AmsterdamUMC | Member page from research data repository (DataverseNL) | |
| Rigshospitalet | https://www.rigshospitalet.dk/maryelizabethshospital/center-for-data-og-effektforskning/om-centeret/Sider/samarbejdsmuligheder.aspx | Data request information from Mary Elizabeths Hospital | Mary Elizabeths Hospital is a part of Rigshospitalet<br><br>In Danish |
| | https://www.regionh.dk/til-fagfolk/forskning-og-innovation/jura-og-data/sider/skabeloner.aspx | Page of agreement templates from Region Hovedstaden | Region Hovedstaden is the regional health authority under which Rigshospitalet operates.<br><br>Contains DTA - Data Transfer Agreement and Databehandleraftale (engelsk) (Data Processing Agreement in English)<br><br>In Danish |
| Erasmus Universitair Medisch | https://www.erasmusmc.nl/en/research/open-and-responsible-science | Page of open science from Erasmus UMC | |
| | https://dtz.erasmusmc.nl/docs/Data_Tran | Generic data transfer agreement | |

| | | | |
|---|---|---|---|
| Centrum Rotterdam | sfer_Agreement.docx | used by Erasmus UMC | |
| | https://dataverse.nl/dataverse/ErasmusMC | Member page from research data repository (DataverseNL) | |
| Medical University of Vienna | https://www.meduniwien.ac.at/web/rechtliches/policy-fuer-forschungsdatenmanagement/ or https://www.meduniwien.ac.at/web/fileadmin/content/serviceeinrichtungen/itsc/it4science/Policy_for_Research_Data_Management.pdf | Research data management policy from MedUniWien | |
| Oslo University Hospital HF | https://ega.uio.no/OUH_specific_dac_guide.html | Data access guideline from Oslo Universitetssykehus | |
| | https://www.ous-research.no/ous/docs/Research-policy-documents/Guideline_research_ethics_and_integrity_OUS.pdf | Research integrity guideline from Oslo Universitetssykehus | |
| | https://ehandboken.ous-hf.no/document/130763 | Data sharing handbook from Oslo Universitetssykehus | In Norwegian |
| | https://ehandboken.ous-hf.no/document/112192 | Health and personal data anonymisation handbook from Oslo Universitetssykehus | In Norwegian |
| | https://ega.uio.no/docs/DTA_template_FEGA_Norway_08.01.20.pdf | Generic data transfer agreement from Oslo Universitetssykehus | |
| | https://ega.uio.no/docs/DPA-between-DAC-and-DataRequestor-template.pdf | Data processor agreement from Oslo Universitetssykehus | |
| | https://www.ous-research.no/ous/docs/Research-policy-documents/Data%20Manage | Data management plan from Oslo Universitetssykehus | |

| | | | |
|---|---|---|---|
| | ment%20Plan%20for%20Reseachers.pdf | | |
| Universitair Medisch Centrum Groningen | https://researchcode.umcgresearch.org/en/w/handling-research-data | Data management code from Groningen UMC | |
| Stichting Radboud universitair medisch centrum | https://www.radboudumc.nl/en/research/open-science/domains/fair-data-and-open-data | Page of open science from Radboud UMC | |
| | https://data.ru.nl/ | Data repository from Radboud Universiteit | |
| | https://data.ru.nl/files/dua-pdfs/RUMC-RA-DUA-1.0.pdf | Data sharing agreement template from Raduboud Universiteit | |
| UZ Leuven | https://gbiomed.kuleuven.be/english/ctc/sub-pages-under-public-homepage/uz-leuvens-policy-regarding-source-data-medical-record-sharing/view | Data sharing policy from UZ Leuven | |
| | https://gbiomed.kuleuven.be/english/ctc/cr-public-website-other-docs/access-request-form-for-3rd-party-access-to-emr | Data access request portal from UZ Leuven | |
| | https://gbiomed.kuleuven.be/english/ctc/sub-pages-under-public-homepage/confidentiality-in-view-of-site-feasibility-activities/view | Page of agreement templates from UZ Leuven | Contains Pharma.be confidentiality agreement and Master confidentiality agreement |
| Unicancer | https://www.unicancer.fr/en/patients-area/personal-data-protection-at-unicancer/ | Patient-facing personal data protection page from Unicancer | Hinted that clinical trial data could be shared with external parties |
| | https://weshare.unicancer.com/ | National digital research infrastructure (WeShare) led by | Currently being deployed and expanded |

| | | | |
|---|---|---|---|
| | | Unicancer | |
| | https://weshare.unicancer.com/studies-and-functionalities/ | WeShare study catalogue | |
| | https://weshare.unicancer.com/wp-content/uploads/2023/04/weshare-infrastructure-operating-charter_v1.0_20230330-3.pdf | WeShare Infrastructure Operating Charter | |
| Karolinska University Hospital | No material found | | |
| Het Nederlands Kanker Instituut | https://www.healthdataspaceamsterdam.nl/en | Regional healthcare data infrastructure joined by the NKI | |
| Odense University Hospital | https://en.ouh.dk/research-and-innovation/research-collaboration | Page of research collaboration from OUH | Hinted that data sharing made possible via OPEN |
| | https://open.rsyd.dk/ | Regional healthcare research support unit that covered OUH | Provided research project management service, data management and storage, workshops and courses<br><br>Version in Danish was more comprehensive |
| | https://www.sdu.dk/da/om-sdu/ledelse-administration/sdurio | Homepage of research development unit from University of Southern Denmark | SDU RIO helped set up data transfer agreement for OUH |
| | https://www.openaire.eu/rdm-handbook<br>or<br>https://dmp.deic.dk/ | Research data management handbook and DeiC roadmap that OUH adopted | |
| Institut Gustave Roussy | https://www.gustaveroussy.fr/fr/recueil-et-utilisation-des-donnees-des-patients-au-sei | Patient-facing personal data protection page from Gustave | Hinted that clinical trial data could be shared with external |

| | | | |
|---|---|---|---|
| | n-de-dr-warehouse | Roussy | parties via Dr WAREHOUSE<br><br>In French |
| | https://weshare.unicancer.com/ | National digital research infrastructure (WeShare) led by Unicancer | Currently being deployed and expanded |
| Region Midtjylland | https://www.fagperson.sundhed.rm.dk/forskning-og-innovation/brug-af-persondata/ | Healthcare-professional-facing page about personal data in research from Region Midtjylland | In Norwegian |
| | https://www.fagperson.hospitalsenhedmidt.dk/afdelinger/regionshospitalet-hammel-neurocenter/university-research-clinic/relink/ | Page of a team (reLINK) from one daughter hospital from Region Midtjylland | Hinted utilising clinical data for research<br><br>In Norwegian |
| | https://www.fagperson.sundhed.rm.dk/forskning-og-innovation/brug-af-persondata/jeg-skal-slette-anonymisere-eller-arkivere-data/ | Page of data anonymisation from Region Midtjylland | In Norwegian |
| | https://www.fagperson.sundhed.rm.dk/forskning-og-innovation/brug-af-persondata/jeg-skal-videregive-data/databehandleraftaler/ | Page of data user agreement from Region Midtjylland | In Norwegian |
| Centre Hospitalier Universitaire de Nantes | No material found | | |
| Universitair Medisch Centrum Utrecht | www.umcutrecht.nl/en/research-data-umc-utrecht | Page of open science from Utrecht UMC | |
| | https://assets-eu-01.kc-usercontent.com/546dd520-97db-01b7-154d-79bb6d | Data sharing guidelines from Utrecht UMC | |

| | | | |
|---|---|---|---|
| | 950a2d/f846a418-d9fd-4f24-bcd1-83c3f555ef2b/Data%20Sharing%20Guidelines%20UMC%20Utrecht.pdf | | |
| | https://dataverse.nl/dataverse/UMCU | Member page from research data repository (DataverseNL) | |
| | https://www.umcutrecht.nl/en/data-request-form-umc-utrecht | Data request form from Utrecht UMC | |
| | https://elsi.health-ri.nl/sites/elsi/files/2020-07/20.30815%20OenO%20Template%20-%20Data%20Sharing%20Agreement.pdf | Nationally-used data sharing agreement template | |
| | https://elsi.health-ri.nl/categorieen/verzamelen-en-uitgeven-van-data-en-lichaamsmateriaal/waar-vind-ik-een-voorbeeld-van-0 | Page on data transfer agreement from ELSI | In Dutch |
| | https://www.uu.nl/en/research/research-data-management/tools-services/tool-to-create-your-dmp-online | Research data management support page on creating DMP from Utrecht University | |
| Hospices Civils de Lyon | No material found | | |
| Region Västra Götaland | No material found | | |
| Ospedale San Raffaele S.r.l. | https://ordr.hsr.it/about | Open Research Data Repository from OSR | |
| | https://research.hsr.it/static/upload/202/20250326_obblighi_open_science.pdf | Slides of open science from OSR | |
| | https://research.hsr.it/static/upload/rio/rio001-0researchintegrityguidelines.pdf | Research integrity guidelines from OSR | Contains research data management guidelines |
