## Supplementary material for "Data sharing policies, requirements, and support from public and private clinical trial sponsors: a survey on top sponsors of clinical trials in Europe": List of included sponsors

Supplementary table 1 - List of included sponsors

| **Canonical name of sponsors** | **Combined count of trials registered** | **Names found on CTIS** |
|---|---|---|
| **Private sponsors** | | |
| Merck Sharp & Dohme LLC | 271 | merck sharp dohme corp,<br>merck sharp dohme llc,<br>merck healthcare kgaa |
| AstraZeneca AB | 230 | astrazeneca ab,<br>astrazeneca farmaceutica spain sa |
| Novartis Pharma AG | 178 | novartis farma spa,<br>novartis pharma ag,<br>novartis pharma gmbh,<br>novartis pharma sas,<br>novartis pharma services ag |
| Janssen-Cilag International | 175 | janssen cilag international,<br>janssen cilag international,<br>johnson johnson enterprise innovation inc,<br>johnson and johnson enterprise innovation inc,<br>janssen vaccines prevention bv |
| F. Hoffmann-La Roche | 171 | f hoffmannla roche ag |
| Boehringer Ingelheim International GmbH | 125 | boehringer ingelheim international gmbh |
| Pfizer Inc | 113 | pfizer inc |
| AbbVie Deutschland GmbH & Co KG | 112 | abbvie deutschland gmbh co kg |
| Eli Lilly & Co | 97 | eli lilly co,<br>eli lilly cork limited |
| Sanofi-Aventis Recherche & Developpement | 96 | sanofiaventis recherche developpement,<br>sanofiaventis research development,<br>sanofi pasteur,<br>sanofi pasteur inc,<br>sanofi aatd inc,<br>sanofi aventis sa,<br>sanofi winthrop industrie |

| | | |
|---|---|---|
| GlaxoSmithKline Research & Development Limited | 89 | glaxosmithkline research development limited,<br>glaxosmithkline biologicals |
| Bristol-Myers Squibb International Corp | 79 | bristol myers squibb international corporation,<br>bristolmyers squibb international corporation,<br>bristolmyers squibb services unlimited company |
| Bayer AG | 62 | bayer ag,<br>bayer consumer care ag |
| Regeneron Pharmaceuticals Inc | 60 | regeneron pharmaceuticals inc |
| Novo Nordisk AS | 52 | novo nordisk as |
| Incyte Corp | 48 | incyte biosciences international sarl,<br>incyte corp |
| Takeda Development Center Americas Inc | 47 | takeda development center americas inc |
| Gilead Sciences Inc | 38 | gilead sciences inc |
| Daiichi Sankyo Co | 32 | daiichi sankyo co ltd,<br>daiichi sankyo inc |
| Sun Pharmaceutical Industries Limited | 31 | sun pharmaceutical industries limited |
| **Public sponsors** | | |
| Assistance Publique - Hôpitaux de Paris | 206 | assistance publique hopitaux de paris |
| Amsterdam UMC | 118 | amsterdam umc,<br>amsterdam umc stichting,<br>stichting amsterdam umc |
| Rigshospitalet | 85 | rigshospitalet |
| Erasmus Universitair Medisch Centrum Rotterdam | 78 | erasmus universitair medisch centrum rotterdam erasmus mc |
| Medical University of Vienna | 72 | medical university of vienna |
| Oslo University Hospital HF | 65 | oslo university hospital hf |
| Universitair Medisch Centrum | 59 | universitair medisch centrum groningen, |

| | | |
|---|---|---|
| Groningen | | university medical center groningen |
| Stichting Radboud universitair medisch centrum | 58 | radboud universitair medisch centrum radboudumc,<br>radboud universitair medisch centrum stichting,<br>stichting radboud universitair medisch centrum,<br>stichting radboud university medical center |
| UZ Leuven | 57 | uz leuven |
| Unicancer | 50 | unicancer |
| Karolinska University Hospital | 49 | karolinska university hospital |
| Universitair Medisch Centrum Utrecht | 47 | universitair medisch centrum utrecht,<br>university medical center utrecht |
| Het Nederlands Kanker Instituut | 45 | het nederlands kanker instituutantoni van leeuwenhoek ziekenhuis stichting,<br>stichting het nederlands kanker instituutantoni van leeuwenhoek ziekenhuis |
| Odense University Hospital | 41 | odense university hospital |
| Institut Gustave Roussy | 41 | institut gustave roussy |
| Region Midtjylland | 40 | region midtjylland |
| Centre Hospitalier Universitaire de Nantes | 39 | centre hospitalier universitaire de nantes |
| Hospices Civils de Lyon | 33 | hospices civils de lyon |
| Region Västra Götaland | 32 | vaestra goetalandsregionen |
| Ospedale San Raffaele S.r.l. | 29 | ospedale san raffaele srl |

| | | |
|---|---|---|
| Celgene Corp | 53 | celgene corp,<br>celgene international ii sarl |

Celgene Corp was acquired by Bristol-Myers Squibb in 2019. Publicly available data sharing policy was since then no longer available. Despite its great amount of registered trials on CTIS, we decided to exclude it and replace it with the next top sponsor.

| | | |
|---|---|---|
| Leiden University Medical Center | 52 | leids universitair medisch centrum lumc,<br>academisch ziekenhuis leiden, |

| | | |
|---|---|---|
| | | leiden university medical center |

A naming harmonisation error in CTIS data led to the exclusion of this public sponsor from the final top-20 list.
